# Racial and ethnic differences in congenital syphilis: mathematical modeling study analyzing the role of prenatal care

**DOI:** 10.64898/2026.01.30.26345236

**Authors:** Minttu M. Rönn, Yizhi Liang, Michelle Bronsard, Ranell L. Myles, Terrika Barham, Harrell W. Chesson, Kathryn Miele, Adrienne Sabety, Lauren Molotnikov, Katherine Hsu, Thomas L. Gift, Joshua A. Salomon

**Affiliations:** Department of Global Health and Population, Harvard T.H. Chan School of Public Health, Boston, MA, USA; Department of Health Policy, Stanford University School of Medicine, Stanford, CA; Office of Public Health Impact, National Center for HIV, Viral Hepatitis, STD, and TB Prevention, Centers for Disease Control and Prevention, Atlanta, GA, USA; Division of STD Prevention, Centers for Disease Control and Prevention, Atlanta, GA, USA; Sexually Transmitted Disease Prevention & HIV/AIDS Surveillance, Massachusetts Department of Public Health, Boston, MA, USA; Section of Pediatric Infectious Disease, Boston Medical Center, Boston, MA, USA

## Abstract

**Background:** Syphilis screening and treatment coverage remain lower than recommended among pregnant women in the US.

**Objective:** We estimated the prevalence and incidence of syphilis among pregnant women, incidence of congenital syphilis and the impact of improved prenatal care cascade by race and ethnicity.

**Design:** Compartmental mathematical model of syphilis natural history, prenatal care, and syphilis screening and testing

**Setting:** United States

**Participants:** Pregnant women

**Measurements:** Women were stratified by race and ethnicity and whether they received any prenatal care. The model was calibrated to epidemiological data for 2019.

**Interventions:** We evaluated improvements among women with any prenatal care and in all pregnant women, and we examined 100% treatment completion, first-trimester testing, first-trimester testing with 100% treatment, testing twice and testing twice with 100% treatment.

**Results:** We estimated that, per 100,000 pregnant women, 110 (95% uncertainty interval [UI] 110-120) had syphilis at pregnancy onset, 13 (95% UI 10-15) acquired syphilis during pregnancy, and 13 (95% UI 12-14) had a syphilis-attributable stillbirth (95% UI 12-14). We estimated a 61% (95% UI 60-62) reduction in non-stillbirth-related congenital syphilis outcomes when at least two syphilis tests were provided and treatment was completed among women who receive prenatal care, and a 98% (95%UI 98- 99%) reduction if two tests were provided and treatment was completed for all pregnant women. The largest benefit of expanding testing and treatment to all pregnant women was seen in non-Hispanic Black and Hispanic populations.

**Limitations:** Results represent national averages and do not account regional variation.

**Conclusion:** Reaching women without prenatal care would substantially reduce congenital syphilis and racial and ethnic differences in congenital syphilis burden.

**Primary Funding source:** Centers for Disease Control and Prevention.

## Introduction

Congenital syphilis is acquired by a fetus when syphilis is untreated or inadequately treated during pregnancy and can cause miscarriage, stillbirth, and neonatal death, as well as lifelong health problems in infected infants who survive.^1,2^ Untreated syphilis in adults is also associated with long-term morbidity, including vision loss, hearing loss, and cardiovascular problems.^1,3^ In 2023, 3,882 cases of congenital syphilis were reported in the United States, more than 8 times the number reported in 2014.^4^ Nearly 90% of reported US congenital syphilis cases in 2022 had mothers with inadequate syphilis screening or treatment, and almost 40% of congenital syphilis diagnoses were among pregnancies without prenatal care.^5^ Modeling studies have estimated that syphilis screening and treatment during pregnancy can prevent congenital syphilis and improve health outcomes at a reasonable cost in the US.^6,7^

Racial and ethnic differences in syphilis and congenital syphilis have been documented,^4^ reflecting complex socioeconomic factors.^8,9^ Healthcare is a factor that influences health, shaping racial and ethnic differences in congenital syphilis incidence. The care cascade for congenital syphilis prevention involves three distinct components: prenatal care, syphilis testing, and syphilis treatment. Syphilis management is part of standard prenatal care.^10,11^ There is also syphilis management outside of prenatal care, for example, in emergency departments^12^ and as part of outreach efforts to specific populations such as women who use substances or experience housing instability.

The extent to which syphilis contributes to stillbirths in the United States is unknown. Of the reported congenital syphilis cases during 2016-2022, 6.7% were stillbirths.^13^ In claims data from 2013, only 7-9% of women with stillbirths received a syphilis test at the time of stillbirth, with 30-35% of women receiving no testing prenatally or at the time of stillbirth.^14^ Diagnoses likely underestimate the true burden because not all women are tested during pregnancy,, particularly stillbirths attributable to syphilis may be underestimated due to low test coverage. In addition, national surveillance data do not distinguish whether syphilis was acquired prior to or during pregnancy. The timing of infection during pregnancy influences the impact of screening during pregnancy on congenital syphilis outcomes.

The impact of variation in utilization of prenatal care on the incidence of congenital syphilis can be estimated using mathematical modeling. We developed a compartmental cohort model of women who are pregnant, stratified by race and ethnicity. We estimated the number of women who had syphilis at the onset of pregnancy (prevalent syphilis) and the number who acquired syphilis during pregnancy (incident syphilis), the incidence of stillbirths attributable to syphilis, and the incidence of other congenital syphilis outcomes. We estimated the potential reduction in congenital syphilis with increased access to prenatal care and with increased access to syphilis screening and treatment regardless of prenatal care access.

## Methods

### Analytic overview

We developed a model to simulate vertical transmission of syphilis among a cohort of pregnant women in the United States in 2019, as we had national diagnosis data in pregnant women by race and ethnicity and syphilis stage for this year. This population reflects women who were pregnant and had live births. The model also includes syphilis-attributable stillbirths (≥ 20 weeks gestation) to capture these congenital syphilis outcomes, but these were the only stillbirths considered in the model. We calibrated the model to multiple syphilis-related epidemiological data to reflect the current care cascade for congenital syphilis by race and ethnicity. We considered receiving any prenatal care, syphilis testing, and syphilis treatment as key steps in the care cascade for congenital syphilis prevention during pregnancy. Both women with and without access to prenatal care could be tested and treated for syphilis, but those with prenatal care were more likely to receive testing and treatment, primarily within prenatal care settings. The model incorporated multiple data on syphilis testing and treatment coverage, and with the model specification accounting for differential testing and treatment coverage by race and ethnicity and by prenatal care.^2,5,13,15^Testing and treatment coverage remains suboptimal also in women with prenatal care, and this is captured in the model.

We estimated several epidemiological outcomes: syphilis infection in pregnant women, stillbirths attributable to syphilis, and other non-stillbirth-related congenital syphilis outcomes (i.e., syphilis- attributable low birth weight, preterm birth, neonatal death, and symptomatic or asymptomatic congenital syphilis in neonates; Figure 1). The model estimates syphilis infections among pregnant women and congenital syphilis outcomes.

**Figure 1.**
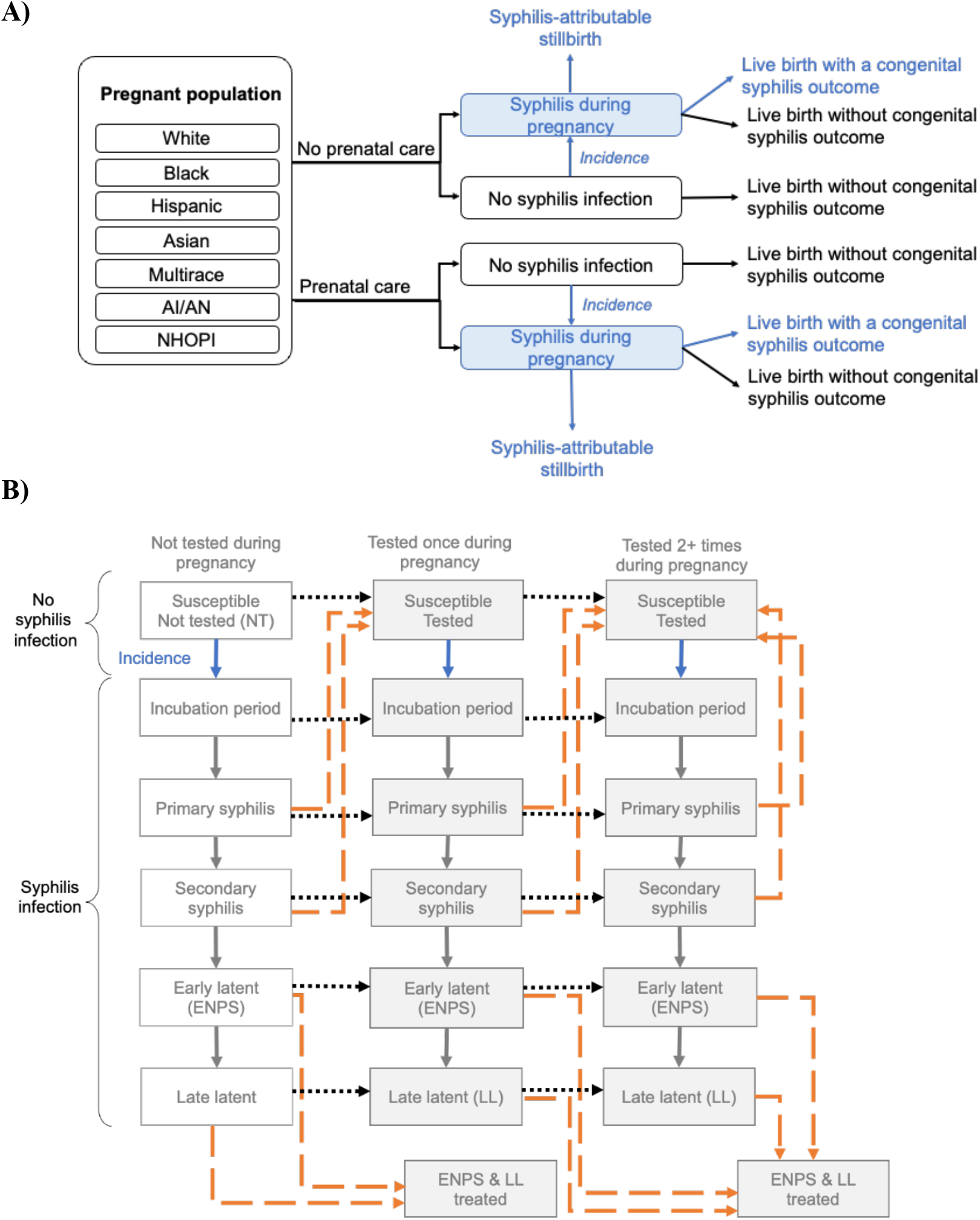
Flowchart of the model structure. **A)** Women who are pregnant by race and ethnicity group*, receipt of ≥1 prenatal care visit, and outcomes after syphilis acquisition, **B)** Natural history, testing, and treatment. Dotted lines in panel B represent testing during pregnancy, with the orange color representing testing combined with successful treatment. *Based on national surveillance congenital syphilis case reporting data, race and ethnicity groups are mutually exclusive. People who were identified as Hispanic were in that category and all other categories include people who are non-Hispanic. Abbreviations: AI/AN: American Indian Alaska Native, NHOPI: Native Hawaiian or Other Pacific Islander, NT: not tested, ENPS: Early non-primary, non-secondary syphilis (clinically known as early latent syphilis), LL: Late latent syphilis

Counterfactual scenarios estimate how closing the gaps in the care cascade for congenital syphilis would impact congenital syphilis outcomes and racial and ethnic differences in these outcomes. These scenarios were informed by national recommendations assuming all populations receive the recommended care for syphilis screening. CDC and the United States Preventive Services Task Force recommend that all women who are pregnant undergo syphilis testing at their initial prenatal visit, and that those at high risk be screened again at 28 weeks and at delivery.^10,11^ The American College of Obstetricians and Gynecologists (ACOG) recommends screening all women who are pregnant at the first visit, and again in the third trimester and at delivery.^16^

### Mathematical model of syphilis in women who are pregnant

A compartmental cohort model (Figure 1) of pregnant women simulates 1) prevalent syphilis at the onset of pregnancy, 2) incident syphilis during pregnancy, 3) any prenatal care utilization, 4) syphilis testing, 5) syphilis treatment, and 6) congenital syphilis outcomes. The model was stratified by race and ethnicity and receipt of any prenatal care. Race and ethnicity categories were based on those in the congenital syphilis national surveillance data: non-Hispanic White (White), non-Hispanic Black (Black), Hispanic, non-Hispanic American Indian or Alaska Native (AI/AN), non-Hispanic Native Hawaiian or Other Pacific Islander (NHOPI), non-Hispanic Asian (Asian), and non-Hispanic Multiracial (Multirace). The modeled health states describe the natural history of syphilis: susceptible and infected, with the latter further divided into stages (incubation period, primary syphilis, secondary syphilis, early non-primary non-secondary syphilis [ENPS; also known clinically as early latent syphilis], and late latent syphilis). The population was also stratified by syphilis testing patterns during pregnancy: not tested, tested once, or tested more than once.

The model was programmed in R and Stan, using difference equations (Supplementary Material). The analytic code is available at: https://github.com/mintturonn/syphilis-incidence-prg.

### Parameterization, calibration and data sources

Key data are summarized in Figure 2, and model parameters are in the Supplementary Material (Table S1). Except for population size and duration of pregnancy, we varied other input parameters over using prior distributions as part of the model calibration. Prior distributions for proportion of women with syphilis at onset of pregnancy (prevalent infection), stage of syphilis among those with a prevalent infection, and weekly probability of acquiring syphilis during pregnancy (incident infection) were varied by race and ethnicity. Syphilis incidence was assumed to be time invariant but varied by race and ethnicity. We assumed those without prenatal care were observed to have higher incidence of congenital syphilis outcomes, lower testing coverage, and lower treatment completion probability, which we reflected in the model parametrization.^2,5,13,15^ We assumed a higher relative risk of syphilis incidence for women without prenatal care compared to those with prenatal care, and this relative risk was assumed to be the same across race and ethnic groups. Prior distributions were varied for durations in each syphilis stage.^17^ Prior distributions for the proportion of women who received prenatal care by race and ethnicity were informed by 2019 birth files from the National Center for Health Statistics, using information from birth certificates for all live births in the United States.^18^ Prior distributions for syphilis testing and treatment were stratified by race and ethnicity and receipt of any prenatal care. Testing coverage and timing of first recorded syphilis test during pregnancy (by trimester) were computed based on testing coverage observed in Medicaid enrolled women who were pregnant, by race and ethnicity, in 2019 (Supplementary Material).^2,5,11,19^ The model therefore captures testing gaps in women without prenatal care and those with prenatal care. We included outcomes related to vertical transmission of syphilis including stillbirths attributable to syphilis and other congenital syphilis outcomes diagnosed at birth. Early pregnancy losses (< 20 weeks gestation), stillbirths due to other reasons (non-syphilis attributable stillbirths), and maternal mortality were not included in the model framework, given the scarcity of relevant data and a need to maintain simplicity in the model framework.

**Figure 2.**
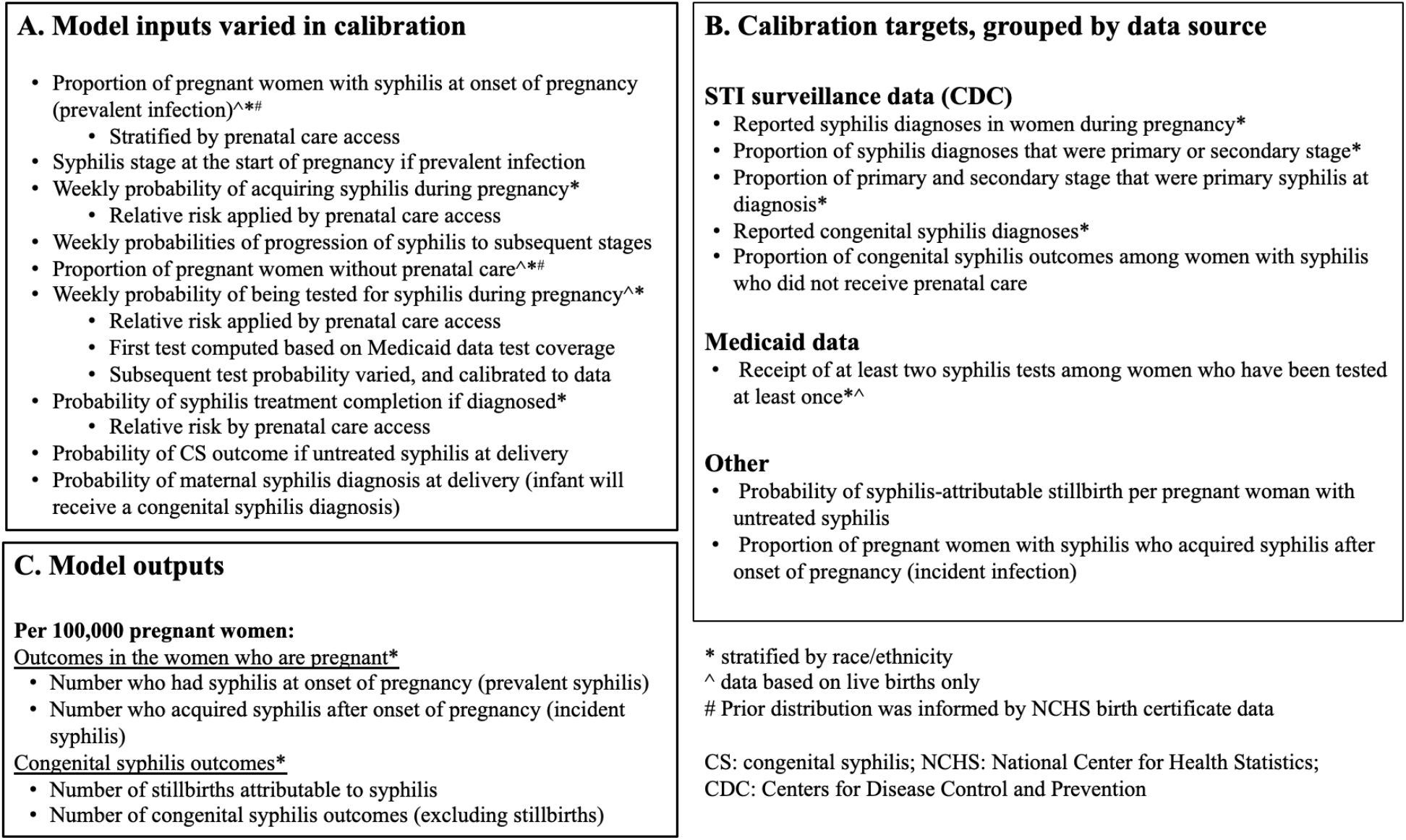
Summary of model inputs (A), key sources for calibration (B), and model outputs (C)

**Figure 2.**
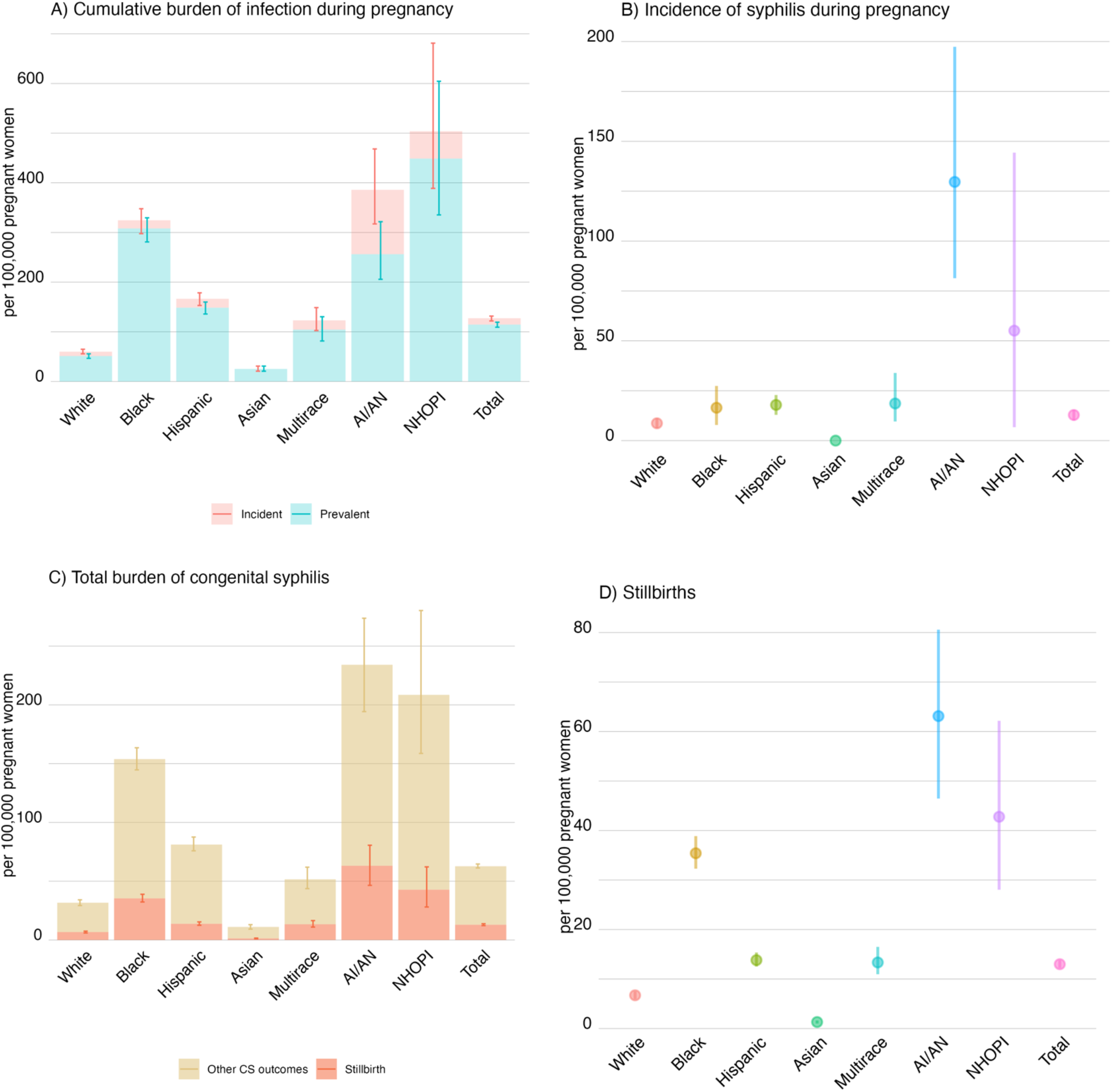
Estimates of syphilis for women who are pregnant and congenital syphilis outcomes by race and ethnicity*. **A)** shows modeled estimates of cumulative burden (syphilis prevalence and incidence) per 100,000 women who are pregnant. **B)** incident syphilis per 100,000 women who are pregnant **C)** stillbirths and other congenital syphilis outcomes and **D)** stillbirths attributable to syphilis. All panels present the median and 95% interval of calibrated model estimates per 100,000 women who are pregnant *) Based on national surveillance congenital syphilis case reporting data 2019, race and ethnicity groups are mutually exclusive. People who were identified as Hispanic were in that category and all other categories include people who are non-Hispanic. Abbreviations: AI/AN: American Indian or Alaska Native, NHOPI: Native Hawaiian or Other Pacific Islander

Prior distribution for syphilis-attributable stillbirths were determined by a weekly probability among 77women who are pregnant with primary syphilis, secondary syphilis, or ENPS; a lower risk was attributed to ENPS based on the association observed between non-treponemal titers and fetal death,^13,19^ Among women who are pregnant with untreated syphilis, there is a pooled 21% probability of stillbirth per pregnancy when the risk of stillbirth due to other reasons is removed. Timing of fetal death was not assessed in all studies, and the pooled estimate might overestimate the rate of stillbirth attributable to syphilis.^20^ Conversely, by assuming no risk of stillbirth from late latent syphilis, despite observed association, we may underestimate stillbirths attributable to this stage of syphilis.^13^

We calibrated the model, using a Markov Chain Monte Carlo (MCMC) algorithm, to the following data targets (Figure 2B), stratified by race and ethnicity unless otherwise stated: i) total syphilis diagnoses in women during pregnancy, ii) proportion of syphilis diagnoses that were primary or secondary stage, and proportion of primary or secondary, which were primary stage, iii) congenital syphilis diagnoses,^21^ iv) proportion of congenital syphilis diagnoses among women with syphilis who did not receive any prenatal care (pooled estimate across race and ethnicity), v) proportion of women receiving at least two syphilis tests (among women who are tested at least once), vi) probability of stillbirth by syphilis treatment status (pooled estimate across race and ethnicity), and vii) proportion of syphilis in pregnant women attributable to incident versus prevalent infection (pooled estimate across race and ethnicity; using diagnosis and staging data from Massachusetts surveillance).

All infants born to women with syphilis at the time of delivery are classified as a non-stillbirth congenital syphilis outcome in the model, though whether they are counted as a congenital syphilis diagnosis depends on whether the syphilis infection in the woman is diagnosed. Most instances of syphilis in pregnant women are diagnosed at or before delivery in the model (parameter prior distribution median 98%, 95%uncertainty interval [95%UI] 96-99%), resulting in both a congenital syphilis outcome and a congenital syphilis diagnosis. When the pregnant woman has undiagnosed syphilis at the time of delivery, the infant is considered to have a congenital syphilis outcome but not a congenital syphilis diagnosis. The assumption was based on less than 2% of congenital syphilis cases receiving a diagnosis after the neonatal period,^22^ which implies there are relatively few undiagnosed vertical transmission events.

The congenital syphilis case definition is “infants whose mother had untreated or inadequately treated syphilis at delivery, regardless of signs in the infant,”^23^ and in the model we assigned a congenital syphilis diagnosis to all infants at delivery, where the mother was diagnosed with syphilis, irrespective of the syphilis stage. Only congenital syphilis diagnoses are calibrated, and the undiagnosed congenital syphilis outcomes remain undetected, but accounted for in the outputs. All diagnoses in women during pregnancy, including those at delivery, were calibrated to 2019 national surveillance data on syphilis diagnoses during pregnancy to ensure we are not overestimating the syphilis burden in women who are pregnant.^23^

### Analysis

We estimated stillbirths and reported other congenital syphilis outcomes as an aggregate measure by race and ethnicity of the pregnant woman.

Across all counterfactual scenarios, we assumed that the initial prevalence and incidence of syphilis by race and ethnicity and prenatal care status remained as in the baseline model, while scenarios differed in the coverage of testing and treatment received during pregnancy. We considered two specifications: in the first, we assumed improvements in the care cascade for congenital syphilis occurred only among those who obtained any prenatal care, while in the second, we assumed improvements occurred among all women who were pregnant, regardless of prenatal care status. In both specifications, we evaluated: (i) 100% treatment completion, (ii) testing at least once during first trimester, (iii) testing at least once during first trimester and 100% treatment completion, (iv) testing at least twice (first and third trimester), and (v) testing at least twice and 100% treatment completion. The scenarios are described in Supplementary Material, Table S2. The scenario of at least two tests during pregnancy with 100% treatment completion best reflects the ACOG recommendation. Whether or not syphilis is diagnosed at delivery does not affect the number of congenital syphilis outcomes in the model, as all women diagnosed at delivery were assigned a congenital syphilis outcome.

We report the median and 95%UI from posterior estimates of the calibrated model. The model assumed all pregnancies were singleton pregnancies (multiple pregnancies such as twins or triplets were not considered).

## Results

The model reproduced the epidemiological data targets as indicated by the calibration fits (Supplementary Material Figures S2-S8).

### Epidemiological estimates

We estimated that per 100,000 women who were pregnant in 2019, 110 (95%UI 110-120) had syphilis at the onset of pregnancy (prevalent infection) and 13 (95%UI 10-15) acquired syphilis during pregnancy (incident infection) (Figure 3). Rates of prevalent and incident syphilis differed markedly by race and ethnicity. The highest prevalence was estimated for the NHOPI (450, 95%UI 340-610), Black (310, 95%UI 280-330), and AIAN (260, 95%UI 210-320) populations. The incidence was highest for the AIAN and NHOPI populations at 130 (95%UI 81-200) and 55 (95%UI 6.7-140), respectively. These outcomes reflect syphilis diagnosis rates that were two and four times higher for women identified as AIAN and NHOPI than the diagnosis rate among all women who were pregnant. They also had higher proportions of primary and secondary syphilis (19% and 13% of all diagnoses, respectively) indicating shorter time between incidence and diagnosis. Most infections in women who were pregnant were estimated to have been acquired prior to pregnancy, accounting for 90% of all infections among women who were pregnant.

**Figure 3.**
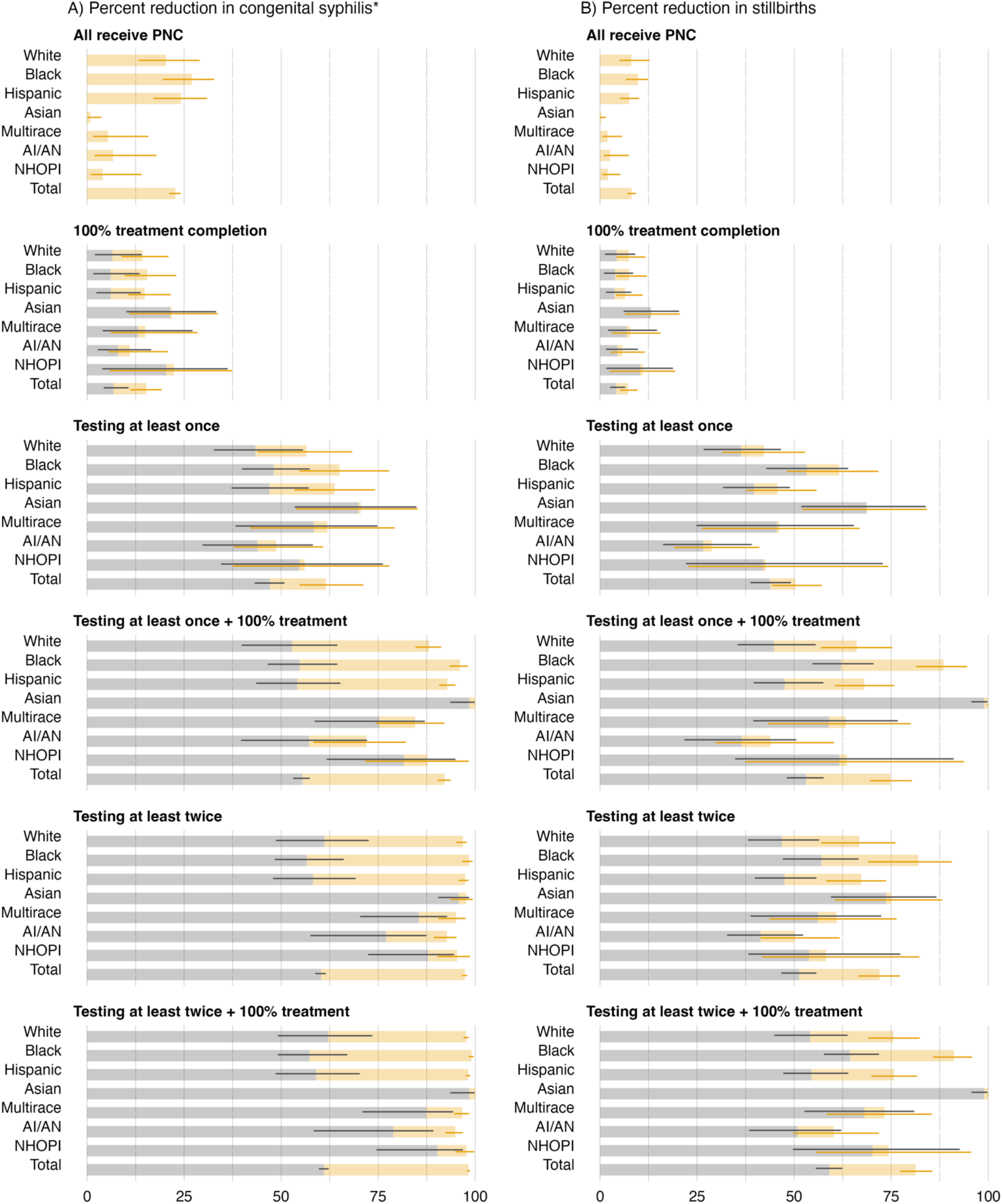
Benefits of increased access to prenatal care by race and ethnicity*: Relative reduction (%) in congenital syphilis by scenario compared to baseline (A) reductions in congenital syphilis outcomes, excluding stillbirths; (B) reductions in stillbirths attributable to syphilis. Grey bars represent the median reduction in congenital syphilis when testing and/or treatment is improved among women receiving any prenatal care (PNC), and orange bars represent the incremental impact if all women receive testing and treatment regardless of PNC status. *) Based on national surveillance congenital syphilis case reporting data 2019, race and ethnicity groups are mutually exclusive. People who were identified as Hispanic were in that category and all other categories include people who are non-Hispanic. Abbreviations: AI/AN: American Indian or Alaska Native, NHOPI: Native Hawaiian or Other Pacific Islander

The rate of congenital syphilis including stillbirths was 63 (95%UI 61-65) per 100,000 pregnant women. The number of syphilis-attributable stillbirths was 5.1 times the reported number in 2019 (480 [450-510] in the model compared to 94 reported) with an incidence of 13 (95%UI 12-14) per 100,000 pregnant women (Figure 2D). NHOPI, Black, and AIAN populations were estimated to have the highest rates of stillbirths attributable to syphilis.

### Impact of prenatal care on congenital syphilis

If women without prenatal care received the same level of syphilis testing and treatment as those receiving any prenatal care, we estimated an 8.1% (95%UI 7.2-9.2%) relative reduction in stillbirths and a 23% (95%UI 21-24%) relative reduction in other congenital syphilis outcomes (Figure 4). The largest relative reductions in other congenital syphilis outcomes were estimated for Black, White, and Hispanic populations with relative reductions of 27% (95%UI 20-33%), 24% (95%UI 17-31%), and 20% (95%UI 13-29%), respectively. This is consistent with lower rates of prenatal care among Black and Hispanic women, and higher prevalence of syphilis among people without prenatal care in these three racial and ethnic groups (Supplementary Material, Figure S10). Similar trends were estimated for stillbirths and other congenital syphilis outcomes, with lower overall reductions in stillbirths.

Asian populations could achieve almost a 100% reduction in congenital syphilis outcomes through increased testing and treatment alone, without increased access to prenatal care. This population had a high rate of prenatal care utilization and a low overall syphilis prevalence and incidence. No other population was estimated to achieve over 90% reduction in congenital syphilis outcomes, unless women with no prenatal care received the same level of testing and treatment as those receiving any prenatal care.

The largest incremental impact of increasing testing and treatment in all women who are pregnant, compared to testing and treatment increasing only among women receiving prenatal care, was estimated in Black and Hispanic populations; non-stillbirth related congenital syphilis outcomes were reduced by an additional 30 percentage points (from approximately 60% to over 90% reduction). Syphilis testing at least twice during pregnancy with 100% treatment completion, if only achieved among women receiving prenatal care, could reduce non-stillbirth related congenital syphilis by 57% (95%UI 49-67%) in the Black population. If the same level of care was extended to women not receiving any prenatal care the reduction was 99% (95%UI 98-100%). The corresponding values for the Hispanic population were 59% (95%UI 49-70) and 98% (95%UI 98-99%). For all pregnant women, we estimated a 61% (95%UI 60-62) reduction in non-stillbirth related congenital syphilis outcomes when only women receiving any prenatal care receive at least two syphilis tests and 100% treatment completion, and 98% (95%UI 98-99%) reduction if we were able to achieve at least two syphilis tests and treatment completion for all pregnant women.

AI/AN and NHOPI populations achieved the highest additional benefits from receiving at least two syphilis tests in comparison to receiving at least one test (during the first trimester). Under the scenarios where all pregnant women receive at least one syphilis test and 100% treatment completion was estimated to reduce non-stillbirth related congenital syphilis outcomes in AIAN populations by 72% (95%UI 58- 82%) while two syphilis tests and 100% treatment completion was estimated to reduce non-stillbirth related congenital syphilis outcomes by 95% (95%UI 92-97)

## Discussion

Improving prenatal care access and providing recommended syphilis screening and treatment are essential to reducing the burden of congenital syphilis. Efforts to increase syphilis screening and treatment need to reach women who do not utilize prenatal care. This can also reduce racial and ethnic differences observed in congenital syphilis, with the largest benefits estimated among Black and Hispanic populations. We estimated that most syphilis among women who are pregnant is acquired before the first week of pregnancy. This emphasizes the importance of an early first syphilis screen during pregnancy, especially given the increased risks of stillbirth in pregnancies complicated by syphilis. AI/AN and NHOPI populations were estimated to have the highest rates incidence of syphilis during pregnancy, and these populations saw the largest relative benefit from testing screening at least twice during pregnancy. The risk of syphilis-attributable stillbirth is highest during early stages of syphilis and, as such, new infections during pregnancy are likely to have a disproportionate impact on stillbirths relative to syphilis acquired prior to pregnancy.

Women without prenatal care face barriers to accessing care and are at increased risk of poor pregnancy outcomes, including congenital syphilis. In Tannis *et al*. women who had a syphilis diagnosis during pregnancy reported histories of incarceration (26.5%), homelessness (47.5%), substance use (63.4%), and receiving medication for opioid use disorder (11.6%).^15^ Partnering with community programs and services that women use can help reach them more effectively outside of traditional prenatal care.^24^ Syphilis testing and treatment in emergency departments and other clinical settings are other important ways to offer syphilis testing and treatment. The recent introduction of rapid point-of-care syphilis tests that facilitate same day treatment offers additional new opportunities to facilitate syphilis management.^12^ Lastly, an important part of the congenital syphilis prevention arsenal is partner testing and treatment.^25^

This analysis examined healthcare as a factor influencing health, inferring its role in maintaining racial and ethnic differences in congenital syphilis via variation in receipt of any prenatal care, syphilis testing, and syphilis treatment. There are distal determinants for these racial and ethnic differences in congenital syphilis. Women from some racial and ethnic minority populations are more likely to be uninsured than White women.^26^ Women with public insurance and underinsured women are more likely to rely on safety-net hospitals, associated with longer wait times and less time per patient.^2,27^ Compared to White women, Black and Hispanic women are less likely to receive adequate prenatal care, with Black women more likely to have four or fewer prenatal visits, and among women who initiate prenatal care, both Hispanic and Black women are more likely to initiate prenatal care in the second or third trimester.^28^

The analysis was based on a model that matched observed differences in syphilis and congenital syphilis, and it reproduced observed syphilis epidemiology, allowing us to estimate stillbirths attributable to syphilis for which limited data exist. U.S. surveillance data suggest the proportion of undiagnosed congenital syphilis is low, with few congenital syphilis cases diagnosed after the neonatal period.^22^ If undiagnosed congenital syphilis remains asymptomatic, the actual proportion could be higher. Some syphilis infections among pregnant women might go undiagnosed without leading to vertical transmission. If undiagnosed infections during pregnancy are underestimated in this study, then the overall burden of syphilis during pregnancy would also be underestimated. As another limitation of the study, we relied largely on data on pregnancies that resulted in a live birth, while syphilis prevalence among women with stillbirths is likely higher than in women with livebirths^29^. Improved estimation of syphilis-attributable stillbirths would require data on the coverage of syphilis prevention among pregnancies that result in a stillbirth. We could not validate our estimate of syphilis-attributable stillbirths due to limited sdata and low syphilis testing coverage when stillbirth occurs.^14^ Our model reflects national averages and does not account for regional differences in syphilis epidemiology. The generalizability of these findings depends on the syphilis prenatal care cascade in a region.^4^

Improving prenatal care access and strengthening syphilis screening and treatment during pregnancy, particularly for populations with no or limited prenatal care, are key in reducing racial and ethnic differences in congenital syphilis in the United States. Ultimately, strengthening the healthcare infrastructure could help to sustain and enhance these improvements over time.

## Data Availability

All data produced in the present work are contained in the manuscript

https://github.com/mintturonn/syphilis-incidence-prg

## Acknowledgments

We would like to thank Elizabeth Torrone for her support in obtaining syphilis surveillance data, and Teresa Puente and Taiwo Abimbola for their guidance during the study.

## Data declarations

Syphilis diagnoses among women who are pregnant by race and ethnicity and stage of syphilis at diagnosis were obtained from the STD Surveillance System, Division of STD Prevention, CDC. These data were determined to be exempt from human subject research. The Medicaid data for this project were accessed using the Stanford Center for Population Health Sciences (PHS) Data Core. The PHS Data Core is supported by a National Institutes of Health National Center for Advancing Translational Science Clinical and Translational Science Award (UL1TR003142) and from Internal Stanford funding. The content is solely the responsibility of the authors and does not necessarily represent the official views of the NIH. All other data are publicly available.

## Notes

**Funding:** This work was supported by the U.S. Centers for Disease Control and Prevention, National Center for HIV, Viral Hepatitis, STD, and TB Prevention Epidemiologic and Economic Modeling Agreement (NU38PS004651).

### Competing Interest Statement

The authors have declared no competing interest.

### Funding Statement

This work was supported by the U.S. Centers for Disease Control and Prevention, National Center for HIV, Viral Hepatitis, STD, and TB Prevention Epidemiologic and Economic Modeling Agreement (NU38PS004651). The findings and conclusions in this report are those of the authors and do not necessarily reflect the official position of the Centers for Disease Control and Prevention.
The PHS Data Core is supported by a National Institutes of Health National Center for Advancing Translational Science Clinical and Translational Science Award (UL1TR003142) and from Internal Stanford funding. The content is solely the responsibility of the authors and does not necessarily represent the official views of the NIH.

### Author Declarations

Syphilis diagnoses among women who are pregnant by race and ethnicity and stage of syphilis at diagnosis were obtained from the STD Surveillance System, Division of STD Prevention, CDC. These data were determined to be exempt from human subject research. The Medicaid data for this project were accessed using the Stanford Center for Population Health Sciences Data Core, and the use of the data are governed by Stanford IRB.

