## Supplemental Material for "Racial and ethnic differences in congenital syphilis: mathematical modeling study analyzing the role of prenatal care"

**Disclaimer:** The findings and conclusions in this report are those of the authors and do not necessarily reflect the official position of the Centers for Disease Control and Prevention.

### Estimation of stillbirth attributable to syphilis

In congenital syphilis surveillance, there were 94 cases of stillbirth reported in 2019 (of 1,870 congenital syphilis diagnoses, or 5% of all congenital syphilis reported). This is most likely an underestimate of the total burden of stillbirth attributable to syphilis. Less than 10% of Medicaid or MarketScan insured women who experienced stillbirth were estimated to have had a syphilis test after a stillbirth, in claims data from 2013, while a higher proportion had evidence of syphilis screening during pregnancy.[1] In an analysis using medical health records in Indiana, 2014-2016, among 35 confirmed stillbirth cases, 51.4% underwent syphilis testing: 31.4% before delivery, 42.9% after, and 22.9% both before and after delivery.[2]

To estimate the burden of stillbirth in pregnant women, we used the syphilis progression in the model together with published estimates of the risk of syphilis during pregnancy. A systematic review estimated that 25.6% (95% CI 18.5-34.2) of women with untreated syphilis experienced stillbirth or fetal loss, while in women without syphilis the estimate was 4.6 (3.0-7.1),[3] and we assumed that the difference (21%) represents the probability of stillbirth attributable to syphilis. The systematic review provides the combined probability of stillbirths and in some cases any fetal loss, which may overestimate the risk of stillbirth attributable to syphilis. The choice between using 25.6% or 21% is not straightforward. In this analysis we aim to capture the burden directly attributable to syphilis which is why we have employed 21%.

These estimates are reported per pregnancy, and we calibrate to this estimate: the denominator (women with untreated syphilis) is defined as women with syphilis infection at the end of their pregnancy plus number of stillbirths, and the numerator is number of stillbirths. In the model, risk of stillbirth begins from 20 pregnancy weeks onwards in women with a syphilis infection if that infection is primary, secondary, or early non-primary non-secondary syphilis (ENPS). During these stages, women experience a weekly probability of stillbirth. This assumes that the risk is dependent on time spent with a syphilis infection, and the risk therefore accumulates over time. If stillbirth occurs, we remove the pregnant woman from the model, and they are no longer counted as a pregnancy.

#### Cohort model structure, parameterization and scenarios specification

**Figure S1.** Schematic overview of the syphilis model. The model is calibrated to observed diagnoses (in dark grey), and screening coverage (grey boxes). Dotted black lines represent testing during pregnancy, and orange lines are testing and succesful treatment for syphilis. See figure footnote for key assumptions.*

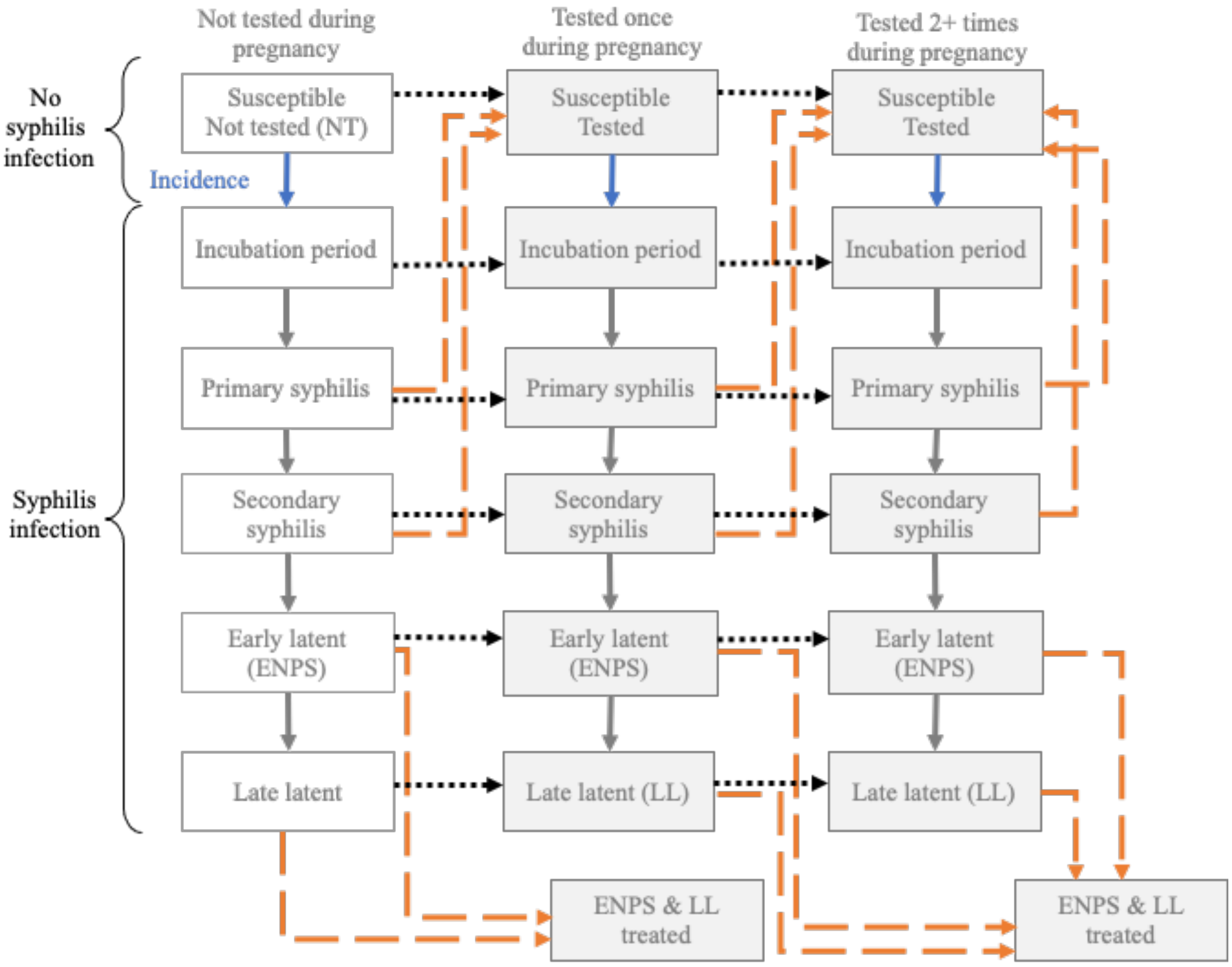

* The sensitivity estimates for syphilis tests and testing algorithms are high[4] and consistent once antibodies have developed (primary syphilis and following stages), and we assume 100% sensitivity given testing. We do not account for false positive tests in the model given that these are very low. Abbreviations: AI/AN: American Indian/Alaska Native, NHOPI: non-Hispanic Native Hawaiian/Other Pacific Islander, NT: not tested, ENPS: Early non-primary, non-secondary syphilis (clinically known as early latent syphilis), LL: Late latent syphilis

**Table S1 Parameters used in the syphilis incidence model.** Prior range shows the median and 95% range from the prior distribution.

| Parameter | Estimate | Prior distribution | Assumptions and considerations |
|---|---|---|---|
| **Pregnant women** | | | |
| **Population size** $N_r$ | | | Birth certificate data: number of pregnant women with live births by race and ethnicity (r).[5]<br><br>Birth certificate data only includes pregnant women with live births. |
| White | 1,912,336 | Fixed | |
| Black | 546,739 | Fixed | |
| AI/AN | 28,440 | Fixed | |
| Asian | 237,928 | Fixed | |
| NHOPI | 9,763 | Fixed | |
| Multiracial | 84,132 | Fixed | |
| Hispanic | 891,210 | Fixed | |
| **Duration of pregnancy (weeks)** | 38 | Fixed | Average duration of pregnancy.[5] |
| **Proportion without documented prenatal care** | | | Birth certificate data on proportion of pregnant women without prenatal care by race and ethnicity.[5] The data were used to define prior distribution based on the proportion and associated 95% uncertainty (based on Wald interval)<br><br>We used rriskDistributions package to define prior ranges based on the proportion and 95% confidence interval of the data point. |
| White | Data: 1.182%<br>Prior range: 1.182%<br>1.167-1.198% | Beta (22077.9882, 1845444.012) | |
| Black | Data: 3.206%<br>Prior range: 3.206%<br>3.159-3.254% | Beta (16915.9679, 510648.032) | |
| AI/AN | Data: 4.104%<br>Prior range: 4.102%<br>3.873-4.341% | Beta (1136.9590, 26569.041) | |
| Asian | Data: 0.932%<br>Prior range: 0.932%<br>0.894-0.972% | Beta (2164.9907, 230078.009) | |
| NHOPI | Data: 5.919%<br>Prior range: 5.915%<br>5.447-6.410% | Beta (545.9408, 8678.059) | |
| Multiracial | Data: 2.167%<br>Prior range: 2.166%<br>2.068-2.268% | Beta (1764.9783, 79692.022) | |
| Hispanic | Data: 2.515%<br>Prior range: 2.515%<br>2.482-2.548% | Beta (21744.9749, 842878.025) | |
| **Syphilis** | | | |
| **Baseline prevalence among women with prenatal care** | | | Syphilis prevalence prior to the first week of pregnancy by receipt of prenatal care.<br><br>We used prevalence of detected infections from birth certificate |
| White | Data: 0.066 %<br>Prior range: 0.033%<br>0.004-0.116% | Beta (1.8425, 4577.7136) | |

| | | | |
|---|---|---|---|
| Black | Data: 0.401 %<br>Prior range: 0.201%<br>0.026-0.697% | Beta (1.8375, 754.9121) | data to inform the prior.[5] We applied an informative prior, which sets the median as approximately 0.5 times the prevalence of detected infections, and 97.5 percentile of the prior as approximately the prevalence of detected infections.<br><br>This set up accounts for detected syphilis burden observed in birth certificate data to be attributed to either pre-pregnancy or acquired during pregnancy. |
| AI/AN | Data: 0.351 %<br>Prior range: 0.176%<br>0.022-0.610% | Beta (1.8382, 863.0222) | |
| Asian | Data: 0.054 %<br>Prior range: 0.027%<br>0.003-0.094% | Beta (1.8427, 5644.4742) | |
| NHOPI | Data: 0.556 %<br>Prior range: 0.278%<br>0.035-0.965% | Beta (1.8353, 543.5985) | |
| Multiracial | Data: 0.186 %<br>Prior range: 0.093%<br>0.012-0.324% | Beta (1.8407, 1632.9726) | |
| Hispanic | Data: 0.165 %<br>Prior range: 0.083%<br>0.011-0.287% | Beta (1.8410, 1842.5158) | |
| **Baseline prevalence among women without prenatal care** | | | |
| White | Data: 0.396 %<br>Prior range: 0.197%<br>0.025-0.687% | Beta (1.8376, 765.8180) | |
| Black | Data: 0.643 %<br>Prior range: 0.322%<br>0.041-1.119% | Beta (1.8340, 469.9397) | |
| AI/AN | Data: 0.887 %<br>Prior range: 0.444%<br>0.056-1.546% | Beta (1.8301, 339.1396) | |
| Asian | Data: 0.046 %<br>Prior range: 0.023%<br>0.003-0.080% | Beta (1.8428, 6587.4169) | |
| NHOPI | Data: 0.368 %<br>Prior range: 0.184%<br>0.024-0.639% | Beta (1.8380, 824.3889) | |
| Multiracial | Data: 0.513 %<br>Prior range: 0.256%<br>0.033-0.890% | Beta (1.8359, 590.1790) | |
| Hispanic | Data: 0.360 %<br>Prior range: 0.180%<br>0.023-0.626% | Beta (1.8382, 842.0016) | |
| **Syphilis stage at the start of pregnancy** | | | To inform the distribution of syphilis stages among those with prevalent infection at the onset of pregnancy, we use the distribution observed among general population of women diagnosed with syphilis. This is reported in the surveillance data as: primary and secondary |
| White | Data:<br>P&S: 0.289, ENPS: 0.276, Late: 0.435 | Dirichlet (22.794, 54.705, 74.146, 116.954) | |
| Black | Data:<br>P&S: 0.240, ENPS: 0.300, Late:0.460 | Dirichlet (23.164, 55.594, 98.366, 150.968) | |

| | | | |
|---|---|---|---|
| AI/AN | Data:<br>P&S: 0.301 ENPS: 0.321, Late: 0.379 | Dirichlet (22.702, 54.485, 82.420, 97.247) | combined, ENPS, late latent, and unknown duration.[6]<br><br>We assume primary and secondary diagnoses are divided between primary and secondary based on the proportional distribution of their respective durations: primary syphilis accounts for 0.29 of diagnoses (45/(45+108) days) and secondary syphilis accounts for 0.71 of diagnoses.[7]<br><br>We assumed that the distribution of syphilis by stage among women with prevalent infection at onset of pregnancy was the same as the distribution of reported syphilis cases by stage among all 15-44-year-old women nationally in 2019. Further, we assumed that this distribution of cases did not vary by prenatal care category. |
| Asian | Data:<br>P&S: 0.141, ENPS: 0.201, Late:0.659 | Dirichlet (23.924, 57.417, 115.955, 380.747) | |
| NHOPI | Data:<br>P&S: 0.223, ENPS: 0.175, Late: 0.602 | Dirichlet (23.292, 55.902, 61.978, 213.480) | |
| Multiracial | Data:<br>P&S: 0.272, ENPS: 0.294, Late: 0.434 | Dirichlet (22.919, 55.007, 84.078, 124.408) | |
| Hispanic | Data:<br>P&S: 0.246, ENPS: 0.168, Late:0.585 | Dirichlet (23.713, 56.912, 117.992, 280.533) | |
| **Incidence during pregnancy,** $\lambda_r$ | 0.067<br>(0.003-0.309) | Beta (1,10) | Weekly probability of new syphilis infections in the susceptible population among those with prenatal care. |
| **Incidence RR for no-PNC** | 1.966<br>(1.480-2.708) | 1+Gamma(10,10) | Multiplier for incidence, used in the population without prenatal care. This is consistent with the higher burden observed in this population. We applied the same incidence RR for all race and ethnicity groups. |
| **Syphilis progression** | | | |
| Weekly probability of progression from incubation period to primary stage, $\sigma_0$ | 0.244 (0.212-0.288) | Beta (137.897 425.818) | Time period in which antibody response is developing.[7,8] We assume syphilis is not detectable by testing during the incubation period. |
| Weekly probability of progression from primary syphilis to secondary, $\sigma_1$ | 0.144 (0.110-0.209) | Beta (48.287, 285.193) | Probability of progressing from primary syphilis to secondary syphilis [7,8] |
| Weekly probability of progression from secondary syphilis to ENPS, $\sigma_2$ | 0.063 (0.049-0.083) | Beta (57.480, 853.096) | Probability of progressing from secondary syphilis to ENPS [7,8] |

| | | | |
|---|---|---|---|
| Weekly probability of progression from ENPS to late, $\sigma_3$ | 0.032 (0.027-0.041) | Beta (112.897, 3352.415) | Probability of progressing from ENPS to late latent syphilis [7,8] |
| **Syphilis testing, weekly probability** $\phi_{r,t,p,w}$ | | | |
| First-time testing $\phi_{r,t=1,p,w}$ | | See description under model equations | First time testing was defined piecewise so that each trimester had a distinct probability of testing. The inputs were defined based on screening coverage among Medicaid insured population. |
| First-time testing RR among women without prenatal care | 0.42 (0.347-0.496) | Beta (71.169, 98.097) | RR for testing probability in the population without prenatal care compared to those with prenatal care. See model equations for implementation. Informed by differences observed in testing among CS cases with and without PNC.[9] |
| Subsequent testing, weekly probability $\phi_{r,t=2,p=2,w=28-38}$ | | | Testing probability among those who receive prenatal care, and who have had one test. This probability is applied for the third trimester only. We used a weekly informative prior and calibrated the parameters to data. We assume that those who did not receive prenatal care could receive only one test during their pregnancy. Calibrated to Medicaid claims on 2+ test coverage data. |
| White | 0.12 (0.029-0.292) | Beta (3, 20) | |
| Black | 0.12 (0.029-0.292) | Beta (3, 20) | |
| AI/AN | 0.12 (0.029-0.292) | Beta (3, 20) | |
| Asian | 0.12 (0.029-0.292) | Beta (3, 20) | |
| NHOPI | 0.12 (0.029-0.292) | Beta (3, 20) | |
| Multiracial | 0.12 (0.029-0.292) | Beta (3, 20) | |
| Hispanic | 0.12 (0.029-0.292) | Beta (3, 20) | |
| **Probability of syphilis treatment completion upon diagnosis,** $\delta_{r,p}$ | | | |
| White | 0.84 (0.650-0.950) | Beta (18.526, 3.865) | In an analysis of 6 states, the proportion of pregnant women diagnosed with syphilis who were adequately treated was on average 57% but varied between 36% to 62% by the race and ethnicity of the population.[10] Given the small sample size (1,476 pregnant women with syphilis) and potential unrepresentativeness of the sample, we defined the same prior for each race and ethnicity group to allow potential variation by race and ethnicity to be modeled. We defined a prior excluding the population whose treatment began <30 days before delivery (n=163) and set the adjusted percentage |
| Black | 0.84 (0.650-0.950) | Beta (18.526, 3.865) | |
| AI/AN | 0.84 (0.650-0.950) | Beta (18.526, 3.865) | |
| Asian | 0.84 (0.650-0.950) | Beta (18.526, 3.865) | |
| NHOPI | 0.84 (0.650-0.950) | Beta (18.526, 3.865) | |
| Multiracial | 0.84 (0.650-0.950) | Beta (18.526, 3.865) | |
| Hispanic | 0.84 (0.650-0.950) | Beta (18.526, 3.865 | |

| | | | |
|---|---|---|---|
| | | | with adequate treatment (65%) as the lower 2.5 percentile and set 95% as the upper 97.5 percentile. This assumes the national average treatment completion is better than estimated in the study. |
| Syphilis treatment completion among women without prenatal care compared to those with prenatal care (RR). | 0.39 (0.18-0.63) | Beta (6.50, 10.05) | In an analysis of 6 states, women without prenatal care had an RR of 0.18 for treatment completion compared to women with timely PNC (12% compared to 68%) and RR of 0.63 for treatment completion compared to women with non-timely PNC (12% vs 20%). We defined these as the 2.5 and 97.5 percentiles of the prior.[10] |
| **Congenital syphilis outcomes** | | | |
| Probability of congenital syphilis outcome at delivery in presence of untreated infection of any stage [excluding stillbirths] $\beta$ | 0.976 (0.959-0.988) | Beta (400,10) | An infant born to any woman identified with a syphilis infection at delivery is considered a probable CS case[11]. This assumes that nearly all women with untreated or inadequately infection at delivery are correctly identified and their newborns diagnosed with CS. |
| Stillbirth attributed to syphilis infection $\psi_s$ | | | Higher non-treponemal titer is associated with worse outcomes[12,13] |
| Weekly probability among those with secondary syphilis from week 20 onwards $\psi_{s=1}$ | 0.018 (0.012-0.025) | Beta (31.901, 1742.210) | Calibrated against data on stillbirth risk among pregnant women. Highest non-treponemal titer is observed during secondary syphilis, and it declines slowly during ENPS. High maternal titer is associated with higher mortality.[12,13] |
| RR1 for probability among those with primary syphilis, i.e. $\psi_{s=2}$=$\psi_{s=1} * RR1$ | 0.853 (0.5-1) | Beta (6.058, 1.300) | RR applied to weekly probability among women with primary syphilis. Among CS cases with lower non-treponemal titer (<=1:4) 82% of deaths were stillbirths.[12] While this does not identify syphilis stage, titer is lower during primary syphilis. In review of stillbirths in the US, relative risk of stillbirth was double with secondary compared to primary syphilis (RR 2.00 (95% CI 1.27– |

| | | | |
|---|---|---|---|
| | | | 3.13)).[13] The difference in risk may also be influenced by the longer duration of secondary stage (almost twice the duration). |
| RR2 for probability among those with ENPS, i.e. $\psi_{s=3} = \psi_{s=1} * RR2$ | 0.25 (0-0.5) | Beta (3.949, 11.196 | Relative risk of stillbirth among women with ENPS was similar to women with primary syphilis (1.10 (95% CI 0.73–1.67)).[13] The duration of ENPS is over 4 times the duration of primary syphilis, and the per week probability would need to be much lower for ENPS than for primary syphilis to generate similar cumulative risk. This presents a simplification given the relationship is unlikely to be constant throughout ENPS (with declining titers). |
| Late syphilis RR3, i.e. $\psi_{s=4} = \psi_{s=1} * RR3$ | 0 | Fixed | Assumed no risk of stillbirth in women with late syphilis |

Abbreviations: AI/AN: American Indian / Alaska Native, CS: congenital syphilis, NHOPI: Native Hawaiian or Pacific Islander, RR: relative risk

**Table S2.** Description of scenarios modeled. **Section A** outlines differences between women without prenatal care (PNC), and women with prenatal care in the calibrated model representing baseline (current) conditions. **Section B** outlines definitions of scenarios used to estimate the benefits achievable if testing and treatment were increased among women with PNC and among women without PNC.

| | **No prenatal care** | **Prenatal care** |
|---|---|---|
| **A. Baseline** | | |
| Population | Proportion reported without PNC informed by birth certificate data. These vary from 1% (Asian) to 6% (NHOPI). | Proportion reported with any PNC informed by birth certificate data. |
| Prevalence, start of pregnancy | Higher prevalence informed by birth certificate data. | Lower prevalence informed by birth certificate data. |
| Incidence during pregnancy | Incidence is assumed to be higher based on overall higher prevalence of detected infections observed in birth certificate data compared to those with prenatal care. | Incidence is assumed to be lower based on overall lower prevalence of detected infections observed in birth certificate data compared to those without prenatal care. |
| Testing | Lower testing coverage informed by literature. Assumed can be tested once only. | Total testing coverage (overall, and coverage of 2+ testing) defined based on data from Medicaid. Higher testing coverage for population receiving PNC. |
| Treatment | Treatment completion lower than for PNC population informed by literature. | Treatment completion less than 100% informed by literature. |
| **B. Counterfactual scenarios** | | |
| **PNC for all** | | |
| All receive PNC level of syphilis prevention | Same level of testing and treatment for women without PNC as to those with PNC at baseline. | Baseline levels of testing and treatment for women with PNC. |
| **100% treatment completeness** | | |
| Baseline PNC coverage | Baseline levels of testing and treatment for women without PNC. | 100% completion of treatment. Testing levels the same as in baseline for women with PNC. |
| All receive PNC level of syphilis prevention | 100% completion of treatment. Testing levels as in baseline for women without PNC. | 100% completion of treatment. Testing levels the same as in baseline for women with PNC. |
| **Testing at least once, during first trimester** | | |
| Baseline PNC coverage | Baseline levels of testing and treatment for women without PNC. | Testing coverage of the first test is increased so that pregnant women with PNC are tested at least once, testing taking place during first trimester. Treatment completion as in baseline for PNC. |
| All receive PNC level of syphilis prevention | Testing coverage of the first test is increased to be 100% (achieved during first trimester) for women without PNC. Treatment completion as in baseline for women without PNC. | Testing coverage of the first test is increased so that pregnant women with PNC are tested at least once, testing taking place during first trimester. Treatment completion as in baseline for PNC. |
| **Testing at least once prior to delivery + 100% treatment** | | |
| Baseline PNC coverage | Baseline levels of testing and treatment for women without PNC. | Testing coverage of the first test is increased so that women with PNC are screened once during first trimester. Treatment completion 100%. |
| All receive PNC level of syphilis prevention | Testing coverage of the first test is increased so that women without PNC are screened once during first trimester. Treatment completion 100%. | Testing coverage of the first test is increased so that women with PNC are screened once during first trimester. Treatment completion 100%. |
| **Testing at least twice prior to delivery** | | |

| | | |
|---|---|---|
| Baseline PNC coverage | Baseline levels of testing and treatment for women without PNC. | Testing coverage is increased so that women with PNC are screened at least twice (first trimester, third trimester). Treatment completion as in baseline. |
| All receive PNC level of syphilis prevention | Testing coverage is increased so that women without PNC are screened at least twice (first trimester, third trimester). Treatment completion as in baseline. | Testing coverage is increased so that women with PNC are screened at least twice (first trimester, third trimester). Treatment completion as in baseline. |
| **Testing at least twice prior to delivery + 100% treatment** | | |
| Baseline PNC coverage | Baseline levels of testing and treatment for women without PNC. | Testing coverage is increased so that women with PNC are screened at least twice (first trimester, third trimester). Treatment completion 100%. |
| All receive PNC level of syphilis prevention | Testing coverage is increased so that women without PNC are screened at least twice (first trimester, third trimester). Treatment completion 100%. | Testing coverage is increased so that women with PNC are screened at least twice (first trimester, third trimester). Treatment completion 100%. |

### Estimating prenatal syphilis screening rates among Medicaid beneficiaries

The rates of prenatal syphilis screening among publicly insured women were obtained using the 2018 and 2019 data from the Transformed Medicaid Statistical Information System Analytic Files in the Medicaid claims database hosted by Stanford Center for Population Health Sciences.[14]

We began by identifying all Medicaid beneficiaries with a delivery and estimating the timing of their pregnancies. Based on Auty et al. (2024), Chen et al. (2023), and a CMS guide on pregnant and postpartum beneficiaries (2023),[15–17] we compiled codes denoting pregnancies and deliveries from Current Procedural Terminology (CPT), Healthcare Common Procedure Coding System (HCPCS), and the tenth revision of the *International Classification of Diseases* (ICD-10) (**Table S3**). Using these codes, we analyzed inpatient and outpatient claims to capture all deliveries each year. We validated the number of deliveries in 2019 by comparing our results to CMS documentation on pregnant and postpartum Medicaid beneficiaries,[18],[5] state-specific vital records and Medicaid coverage data,[18] KFF records of Medicaid-financed births,[19] and 2019 birth certificate data.

We then estimated the timing of each pregnancy by subtracting nine months from the delivery date, consistent with the method used by Lanier et al. (2022) and Hammerslag et al. (2023).[20,21] These studies assume a 9-month (281-day) prenatal period, i.e., that all deliveries occurred at full term.
Among beneficiaries with a delivery, we assessed whether they received a syphilis screening during the 9 months preceding their delivery. Syphilis screening was identified using CPT and HCPCS codes (**Table S3**) denoting a screening or a full obstetric panel. Data was aggregated by race and ethnicity and by whether beneficiaries received one or multiple screenings.

There are several limitations to our approach. Although deliveries that ended in stillbirths are included in our pregnancy count, incomplete pregnancies (e.g., abortions and miscarriages) are not counted, and these represent a non-negligible proportion of pregnancies. In 2019, there were 431,278 abortions, miscarriages, and stillbirths, compared to approximately 1,609,000 live births.[5] Additionally, we did not adjust pregnancy timelines for preterm or late-term deliveries, as gestational age codes in claims data were often missing. We restricted our analysis to one delivery per woman in 2019. Finally, prenatal syphilis screenings performed as part of bundled tests that were not labeled as an obstetric panel were not counted, potentially leading to underreporting of screening rates.

**Table S3.** International Classification of Diseases (ICD-10)

| | File type | Code | Code type |
|---|---|---|---|
| Live birth | Diagnosis | O80, O82, Z370, Z372, Z373, Z375, Z3750, Z3751, Z3752, Z3753, Z3754, Z3759, Z376, Z3760, Z3761, Z3762, Z3763, Z3764, Z3769, Z379, Z390 | ICD-10-CM |
| Live birth | Procedure | 10D00Z0, 10D00Z1, 10D00Z2, 10D07Z3, 10D07Z4, 10D07Z5, 10D07Z6, 10D07Z7, 10D07Z8,10D17ZZ, 10D18ZZ,10E0XZZ, 10S07ZZ, 10A00ZZ, 10A03ZZ,10A04ZZ, 10A07Z6, 10A07ZW, 10A07ZX, 10A07ZZ, 10A08ZZ, 10D27ZZ, 10D28ZZ,10J20ZZ, 10J23ZZ,10J24ZZ, 10J27ZZ,10J28ZZ, 10J2XZZ,10S20ZZ, 10S23ZZ,10S24ZZ, 10S27ZZ,10S28ZZ, 10T20ZZ,10T23ZZ, 10T24ZZ,10T27ZZ,10T28ZZ | ICD-10-PCS |
| Live birth | Procedure | 59400, 59409, 59410, 59510, 59514, 59515, 59610, 59612, 59614, 59618, 59620, 59622 | CPT |
| Syphilis screening | Procedure | 86592, 86593, 86780, 87285, 0064U, 0065U, 0210U | CPT |
| Syphilis screening | Procedure | G9228 | HCPCS |
| Obstetric panel | Procedure | 80055, 80081 | CPT |

### Model equations

Time step used is one week, and the equations represent difference equations.

Susceptible $X_{r,p,t,w}$ stratified by race and ethnicity (r), prenatal care access (p), testing status (t), and time (w). The model was simulated with weekly timesteps for 38 weeks (average duration of pregnancy).

Among those who have not been tested (t=0):

$$X_{r,p,t=0,w} = -\lambda_{r,p} X_{r,p,t=0,w} - \phi_{r,p,t,y} X_{r,p,t=0,w}$$

Among those who have been tested once previously (t=1)

$$X_{r,p,t=1,w} = -\lambda_{r,p} X_{r,p,t=1,w} + \phi_{r,p,t=0,w} X_{r,p,t=0,w} - \phi_{r,p,t=1,w} X_{r,p,t=1,w} + \delta_{r,p} \phi_{r,p,t=0,w} P_{r,p,t=0,w} + \delta_{r,p} \phi_{r,p,t=0,w} S_{r,p,t=0,w}$$

And among those who have been tested at least twice previously (t=2)

$$X_{r,p,t=2,w} = -\lambda_{r,p} X_{r,p,t=1,w} + \phi_{r,p,t=1,w} X_{r,p,t=1,w} + \phi_{r,p,t=1,w} P_{r,p,t=1,w} + \delta_{r,p} \phi_{r,p,t=1,w} S_{r,p,t=1,w} + \delta_{r,p} \phi_{r,p,t=2,w} P_{r,p,t=2,w} + \delta_{r,p} \phi_{r,p,t=2,w} S_{r,p,t=2,w}$$

Where $\lambda_r$ was the weekly probability of incident infection among the susceptible population. This was assumed to be constant through pregnancy, and by testing status, but it was allowed to vary by race and ethnicity.

$\phi_{r,p,t,w}$ is race and ethnicity and time-varying weekly probability of testing for syphilis. We assumed that probability of testing does not depend on syphilis stage; primary and secondary stages are associated with symptoms. This was done given the relatively high coverage of syphilis screening, which assumes that screening probability is at least as high as care seeking due to symptoms. Probability of syphilis treatment completion upon diagnosis was defined by $\delta_{r,p}$.

We allowed probability of first-time testing (t=1) to differ from subsequent testing. The time-varying component was constructed piecewise so that for weeks 1-13 we assigned first trimester (m=1) probability, for weeks 14-27 second trimester (m=2) probability, and weeks 28-38 third trimester (m=3) probability.

We calculated weekly probability of screening by trimester as:

$$\phi_{r,p,t=1,w=0\ldots13} = 1 - (1 - \tau_{rm=1})^{1/14}$$

$$\phi_{r,p,t=1,w=14\ldots27} = 1 - (1 - \tau_{rm=2})^{1/14}$$

$$\phi_{r,p,t=1,w=28\ldots37} = 1 - (1 - \tau_{rm=3})^{1/10}$$

Where $\tau_{rm}$ was the trimester-specific testing coverage (probability of screening during the trimester) calculated as $\tau_{rm} = \frac{Z_{rm}}{F_{rm}}$, where the denominators were defined as number of women who had not been screened by the start of the trimester ($F_{rm}$), and numerators were number of women who were screened for the first time during the trimester ($Z_{rm}$).

We used the population size of pregnant women by race and ethnicity ($N_r$) and screening coverage data along with the distribution of first-time screen by trimester to estimate $F_{rm}$ and $Z_{rm}$:

$F_{rm=1} = N_r$

$F_{rm=2} = N_r - N_r * W_{rm=1}$

$F_{rm=3} = N_r - N_r * W_{rm=1} - N_r * W_{rm=2}$

$W_{rm} = M_r * dM_{rm}$, where $M_r$ was overall screening coverage (screened at least once) by race and ethnicity and $dM_{rm}$ was the proportion of first screen by trimester. See data section on Medicaid screening coverage for the estimates used.

Number screened during each trimester ($Z_{rm}$) was calculated as:

$Z_{rm=1} = N_r * W_{rm=1}$

$Z_{rm=2} = F_{rm=3} - F_{rm=2}$

$Z_{rm=3} = N_r * M_r - N_r * W_{rm=2} - N_r * W_{rm=3}$

Syphilis natural history is modeled with 5 stages:

1) Infected, *incubation stage*, $C_{r,p,t,w}$

$$C_{r,p,t=0,w} = \lambda_{r,p} X_{r,p,t=0,y} - \sigma_0 C_{r,p,t=0,w} - \phi_{r,p,t,w} C_{r,p,t=0,w}$$

$$C_{r,p,t=1,w} = \lambda_{r,p}X_{r,p,t=1,y} - \sigma_0 C_{r,p,t=1,w} + \phi_{r,p,t,w}C_{r,p,t=0,w} - \phi_{r,p,t=1,w}C_{r,p,t=1,w}$$

$$C_{r,p,t=2,w} = \lambda_{r,p}X_{r,p,t=2,y} - \sigma_0 C_{r,p,t=2,w} + \phi_{r,p,t,w}C_{r,p,t=1,w}$$

2) *Primary syphilis*, $P_{r,p,t,w}$

$$P_{r,p,t=0,w} = \sigma_0 C_{r,p,t=0,w} - \sigma_1 P_{r,p,t=0,w} - \phi_{r,t=0,w}P_{r,p,t=0,w} - \psi_{s=1}P_{r,p,t=0,w}$$

$$P_{r,p,t=1,w} = \sigma_0 C_{r,p,t=1,w} - \sigma_1 P_{r,p,t=1,w} - \phi_{r,t=1,w}P_{r,p,t=1,w} - \psi_{s=1}P_{r,p,t=1,w} + \left(1-\delta_{r,p}\right)\phi_{r,t=0,w}P_{r,p,t=0,w}$$

$$P_{r,p,t=2,w} = \sigma_0 C_{r,p,t=2,w} - \sigma_1 P_{r,p,t=2,w} - \phi_{r,t=2,w}P_{r,p,t=2,w} - \psi_{s=1}P_{r,p,t=2,w} + \left(1-\delta_{r,p}\right)\phi_{r,t=1,w}P_{r,p,t=1,w}$$

3) *Secondary syphilis*, $S_{r,t,w}$

$$S_{r,p,t=0,w} = \sigma_1 P_{r,p,t=0,w} - \sigma_2 S_{r,p,t=0,w} - \phi_{r,p,t=0,w}S_{r,p,t=0,w} - \psi_{s=1}S_{r,p,t=0,w}$$

$$S_{r,p,t=1,w} = \sigma_1 P_{r,p,t=1,w} - \sigma_2 S_{r,p,t=1,w} - \phi_{r,p,t=1,w}S_{r,p,t=1,w} - \psi_{s=1}S_{r,p,t=1,w} + \left(1-\delta_{r,p}\right)\phi_{r,t=0,w}S_{r,p,t=0,w}$$

$$S_{r,p,t=2,w} = \sigma_1 P_{r,p,t=2,w} - \sigma_2 S_{r,p,t=2,,w} - \phi_{r,p,t=1,w}S_{r,p,t=2,w} - \psi_{s=1}S_{r,p,t=2,w} + \left(1-\delta_{r,p}\right)\phi_{r,t=1,w}S_{r,p,t=1,w}$$

4) *Early non-primary, non-secondary (ENPS, also known as early latent)* $E_{r,t,w}$

$$E_{r,p,t=0,w} = \sigma_2 S_{r,p,t=0,w} - \sigma_3 E_{r,p,t=0,w} - \phi_{r,p,t=0,w}E_{r,p,t=0,w} - \psi_{s=1}E_{r,p,t=0,w}$$

$$E_{r,p,t=1,w} = \sigma_2 S_{r,p,t=1,w} - \sigma_3 E_{r,p,t=1,w} - \phi_{r,p,t=1,w}E_{r,p,t=1,w} - \psi_{s=1}E_{r,p,t=1,w} + \left(1-\delta_{r,p}\right)\phi_{r,t=0,w}E_{r,p,t=0,w}$$

$$E_{r,p,t=2,w} = \sigma_2 S_{r,p,t=0,w} - \sigma_3 E_{r,p,t=2,w} - \phi_{r,p,t=2,w}E_{r,p,t=2,w} - \psi_{s=1}E_{r,p,t=2,w} + \left(1-\delta_{r,p}\right)\phi_{r,p,t=1,w}E_{r,p,t=1,w}$$

5) *Late latent*, $L_{r,p,t,w}$

$$L_{r,p,t=0,w} = \sigma_3 E_{r,p,t=0,w} - \phi_{r,p,t=0,w}L_{r,p,t=0,w}$$

$$L_{r,p,t=1,w} = \sigma_3 E_{r,p,t=1,w} - \phi_{r,p,t=1,w}L_{r,p,t=1,w} + \left(1-\delta_{r,p}\right)\phi_{r,t=0,w}L_{r,p,t=0,w}$$

$$L_{r,p,t=2,w} = \sigma_3 E_{r,p,t=2,w} - \phi_{r,p,t=2,w}L_{r,p,t=2,w} + \left(1-\delta_{r,p}\right)\phi_{r,p,t=1,w}L_{r,p,t=1,w}$$

Progression of untreated syphilis was defined by $\sigma_0$ (weekly probability of transition from incubation period to primary syphilis), $\sigma_1$ (weekly probability of primary syphilis progressing to secondary syphilis),

$\sigma_2$ (weekly probability of secondary syphilis progressing to ENPS syphilis), and $\sigma_3$ (weekly probability of ENPS syphilis progressing to late syphilis). We assumed that women do not spontaneously clear infection, and that women with syphilis will remain infected unless diagnosed and treated. Syphilis attributable stillbirths were modeled as a weekly probability $\psi_s$ from week 20 onwards.

For women who are treated during primary and secondary stage, we assumed they return to susceptible state with no immunity acquired, while for women treated during ENPS or late latent stages, we assumed that there is sufficient temporary immunity to protect against new infection during the same pregnancy. This was included in the model with a treated compartment:

6) Treated during ENPS or late latent stage

   Tested once (t=1)

$$T_{r,p,t=1,w} = \phi_{r,p,t=0,w}E_{r,p,t=0,w} + \phi_{r,p,t=0,w}L_{r,p,t=0,w}$$

   Tested more than once (t=2)

$$T_{r,p,t=2,w} = \phi_{r,p,t=1,w}E_{r,p,t=1,w} + \phi_{r,p,t=1,w}L_{r,p,t=1,w} + \phi_{r,p,t=2,w}E_{r,p,t=2,w} + \phi_{r,p,t=2,w}L_{r,p,t=2,w}$$

Modeled syphilis incidence ($i_r$) is calculated as the cumulative number of new infections during pregnancy by race and ethnicity:

$$i_r = \sum_{w,p}(\lambda_{r,p}X_{r,p,t=0,w} + \lambda_{r,p}X_{r,p,t=1,w} + \lambda_{r,p}X_{r,p,t=2,w})$$

Modeled diagnoses are calculated as cumulative number of diagnoses during pregnancy by stage at time of diagnosis ($p_r, s_r, e_r, l_r$), and by race and ethnicity of the woman being diagnosed:

$$p_r = \sum_{p,t,w}\phi_{r,p,t,w}P_{r,p,t,w}$$

$$s_r = \sum_{p,t,w}\phi_{r,p,t,w}S_{r,p,t,w}$$

$$e_r = \sum_{p,t,w}\phi_{r,p,t,w}E_{r,p,t,w}$$

$$l_r = \sum_{p,t,w}\phi_{r,p,t,w}L_{r,p,t,w}$$

Total diagnoses are the sum of these.

Modeled congenital syphilis infections at birth by race and ethnicity, by disease stage and by prenatal care access

$$cs\_p_{r,p} = \sum_{t}\beta P_{r,p,t,w=38}$$

$$cs\_s_{r,p} = \sum_t \beta S_{r,p,t,w=38}$$

$$cs\_e_{r,p} = \sum_t \beta E_{r,p,t,w=38}$$

$$cs\_l_{r,p} = \sum_t \beta L_{r,p,t,w=38}$$

Modeled stillbirths attributable to syphilis by race and ethnicity

$$sb\_p_{r,p} = \sum_t \psi_s P_{r,p,t,w=38}$$

$$sb\_s_{r,p} = \sum_t \psi_s S_{r,p,t,w=38}$$

$$sb\_e_{r,p} = \sum_t \psi_s E_{r,p,t,w=38}$$

$$sb\_l_{r,p} = \sum_t \psi_s L_{r,p,t,w=38}$$

### Calibration and calibration data

Syphilis prevalence at the start of pregnancy was informed by birth certificate data of syphilis positivity. The prior distributions were set so that the upper end of the prior distribution corresponds to the positivity observed in the birth certificate data. Syphilis stage distribution among women with infection at the start of the pregnancy was informed by the stage distribution among 15-44-year-old women in the general population diagnosed with syphilis. This assumes that the diagnosis distribution in the general population of women reflects the underlying stage distribution in the population with syphilis. Routine syphilis screening recommendations for women who are not pregnant are based on local syphilis prevalence and individual risk factors. This may overestimate the proportion of infections in the primary and secondary stages given these stages are the most symptomatic and more likely to be diagnosed.

The model was developed in Stan[22] which uses Monte Carlo Metropolis Hastings algorithm. We calibrated with 3 chains, each with 9,000 simulations, and collected the last 50 simulations from each chain.

Key data for incidence estimation are diagnoses during pregnancy by each stage of syphilis and by race and ethnicity. These data are not publicly available and were obtained from the Division of STD Prevention at the Centers for Disease Control and Prevention (CDC).

1. We calibrated to observed syphilis diagnoses (D) among pregnant women by race and ethnicity (r) for each syphilis stage (P=primary, S=secondary, E=ENPS, L=late latent). Women with congenital syphilis outcome in a newborn were included in the number of syphilis diagnoses among pregnant women:

$$d_r = p_r + s_r + e_r + l_r + \sum_p cs\_p_{r,p} + \sum_p cs\_s_{r,p} + \sum_p cs\_e_{r,p} + \sum_p cs\_l_{r,p}$$

And $\frac{d_r}{N_r}$ is the model estimated diagnosis probability per population applied to binomial likelihood:

$$D_r \sim Binomial(N_r, \frac{d_r}{N_r})$$

Where the data presents sum of diagnoses by stage: $D_r = DP_r + DS_r + DE_r + DL_r$

2. Syphilis staging during diagnosis is associated with uncertainty, and we calibrated to the stages where symptoms are present (primary and secondary) with the assumption that the accuracy of the staging of these diagnoses is better than for ENPS or late latent syphilis. Using beta likelihood, we calibrated to proportion of diagnoses occurring during primary or secondary stage:

$$\frac{DP_r + DS_r}{D_r} \sim Beta(\alpha_{1_r}, \beta_{1_r})$$

And proportion of primary or secondary diagnoses that are primary:

$$\frac{DP_r}{DP_r + DS_r} \sim Beta(\alpha_{2_r}, \beta_{2_r})$$

Where $\alpha$ and $\beta$ are calculated based on the model estimated proportion of cases observed for the outcome, and variance associated with the data.

3. We calibrated to congenital syphilis diagnoses, $CS\_D_r$ calculating the congenital syphilis probability per population in the model

$$cs\_d_r = \sum_p cs\_p_{r,p} + \sum_p cs\_s_{r,p} + \sum_p cs\_e_{r,p} + \sum_p cs\_l_{r,p}$$

Using binomial likelihood:

$$CS\_D_r \sim Binomial(N_r, \frac{cs\_d_r}{N_r})$$

4. We calibrated to the proportion of congenital syphilis outcomes in women without prenatal care utilization

$$pr_{cs} = \frac{\sum_r cs\_p_{r,p=0} + \sum_r cs\_s_{r,p=0} + \sum_r cs\_e_{r,p=0} + \sum_r cs\_l_{r,p=0}}{\sum_{r,p} cs\_p_{r,p} + \sum_{r,p} cs\_s_{r,p} + \sum_{r,p} cs\_e_{r,p} + \sum_{r,p} cs\_l_{r,p}}$$

We used the data on proportion of CS cases in women without prenatal care, and calculated the expected number of congenital cases 5,000 women (population size decided arbitrarily), 37.9%*5000=1895, used in binomial likelihood:

$$1895 \sim Binomial(5000, pr_{cs})$$

5. We calibrated to the coverage of subsequent screening among women tested once was defined as:

$$scov2_r =$$

$$\frac{\sum_p X_{r,p,t=2,w=38} + C_{r,p,t=2,w=38} + P_{r,p,t=2,w=38} + S_{r,p,t=2,w=38} + E_{r,p,t=2,w=38} + L_{r,p,t=2,w=38} + T_{r,p,t=2,w=38}}{\sum_{p,t=1}^{t=2} X_{r,p,t,w=38} + C_{r,p,t,w=38} + P_{r,p,t,w=38} + S_{r,p,t,w=38} + E_{r,p,t,w=38} + L_{r,p,t,w=38} + T_{r,p,t,w=38}}$$

The overall screening coverage (tested at least once at any point during pregnancy) was defined so that it corresponded to data. Data were taken from an analysis of Medicaid insured population in the US using national syphilis testing coverage at national-level, and for all seven racial and ethnic groups included in the model.

Overall subsequent screening coverage is defined as $SCOV2_r$;

$SCOV2_r \sim Binomial(N2_r, scov2_r)$

Where $N2 = N_r * (1 - SCOV1_r)$ with $SCOV1_r$

6. We calibrated the modeled syphilis attributable stillbirths to the estimated of stillbirth risk among women with untreated syphilis: the denominator (women with untreated syphilis) is defined as women with syphilis infection at the end of their pregnancy plus number of stillbirths, and the numerator is number of stillbirths.

$$pr_{sb} = \frac{\sum_r sb\_p_{r,p=0} + \sum_r sb\_s_{r,p=0} + \sum_r sb\_e_{r,p=0} + \sum_r sb\_l_{r,p=0}}{\sum_{r,p,t} P_{r,p,t,w=38} + \sum_{r,p,t} S_{r,p,t,w=38} + \sum_{r,p,t} E_{r,p,t,w=38} + \sum_{r,p,t} L_{r,p,t,w=38}}$$

We calculated the expected number of stillbirths for 5,000 women with untreated syphilis (population size decided arbitrarily), 21%*5000=1050, used in binomial likelihood:

$1050 \sim Binomial(5000, pr_{sb})$

7. We calibrated the proportion of syphilis infections that were incident: $\frac{model\_Incid}{model\_incid + model\_prevalent}$. Syphilis diagnoses among pregnant women in Massachusetts in 2022 were used to estimate proportion of diagnoses that could have been due to incident infection based on staging of infection and duration of pregnancy at diagnosis, as well as additional infection available from previous tests when available, defined as $\frac{D\_Incid}{D\_Total}$). The estimate is based on 310 diagnoses, and we set this as $D\_Total$ used in the binomial likelihood:

$D\_Incid \sim Binomial(D\_Total, \frac{model\_Incid}{model\_incid + model\_prevalent})$

### Calibration results

**Figure S2.** Calibration (A) and comparison (B) to syphilis diagnoses in pregnant women in 2019 in the United States. Surveillance data.

### S2A

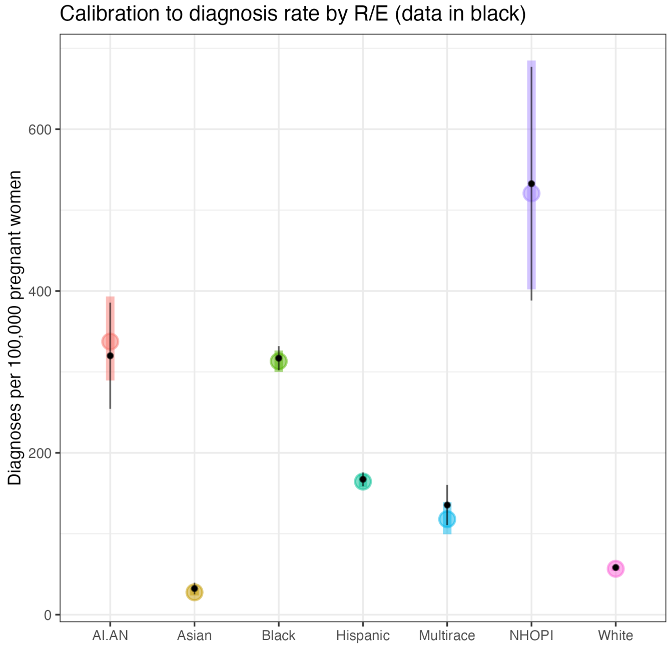

### S2B

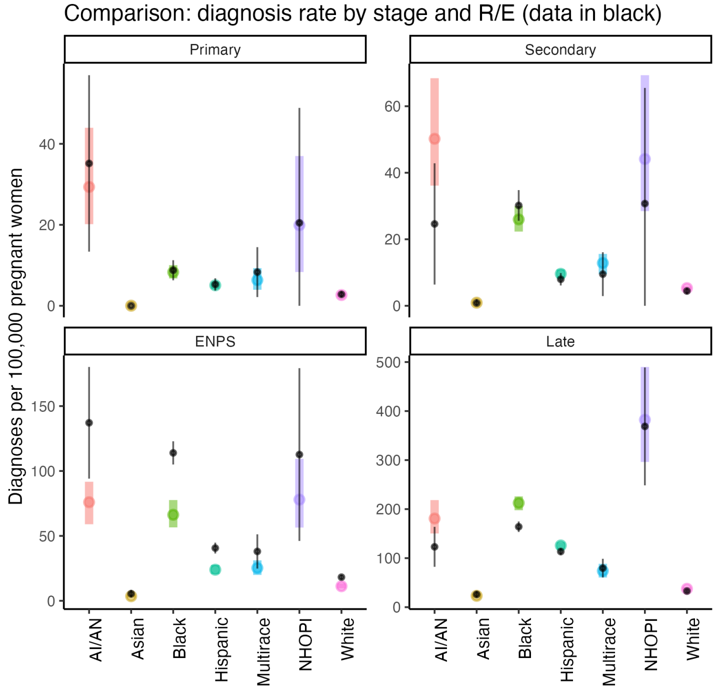

**Figure S3.** Calibration to proportion of diagnoses that are primary or secondary syphilis (of total diagnoses), and proportion of diagnoses that are primary syphilis (of primary and secondary) in 2019 in the United States. Surveillance data.

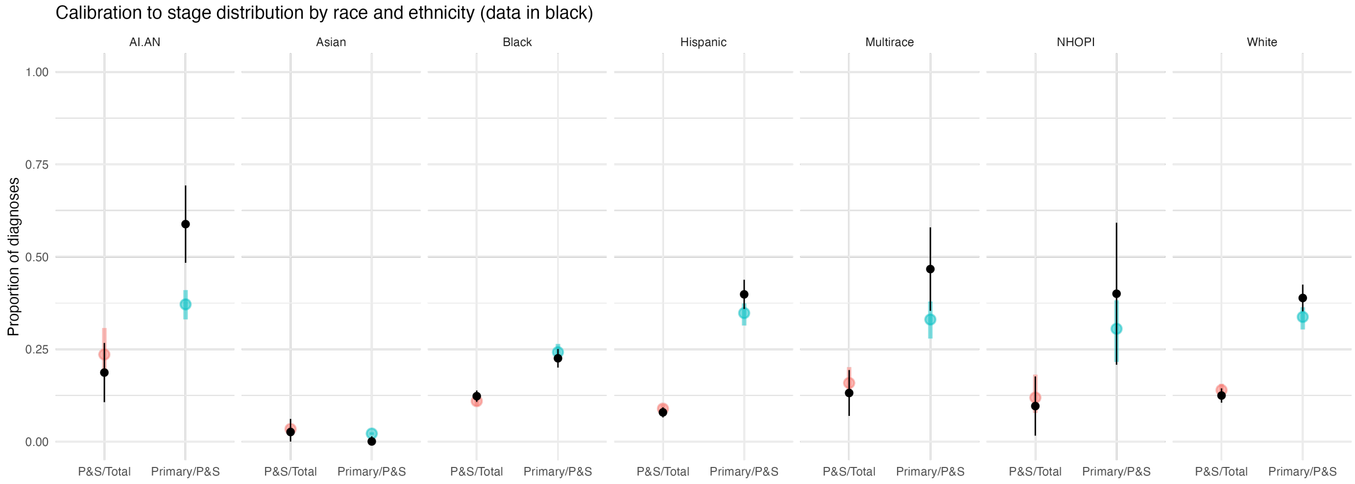

**Figure S4.** Calibration (A) and comparison of input data (B) against overall screening coverage estimated in the model in pregnant women in 2019 in the United States. Data using all Medicaid claims in 2019.

**S4A**

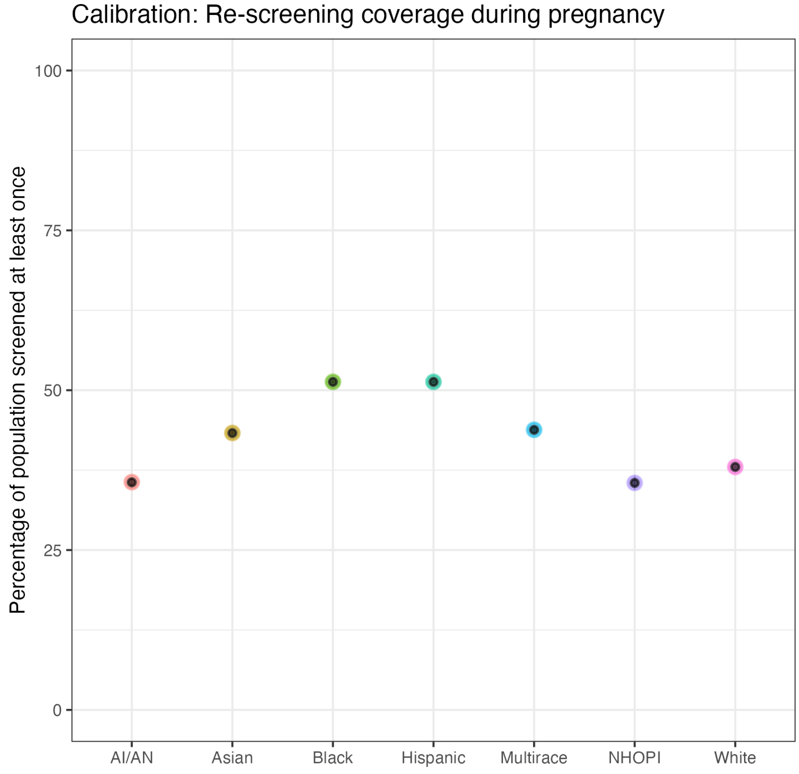

**S4B**

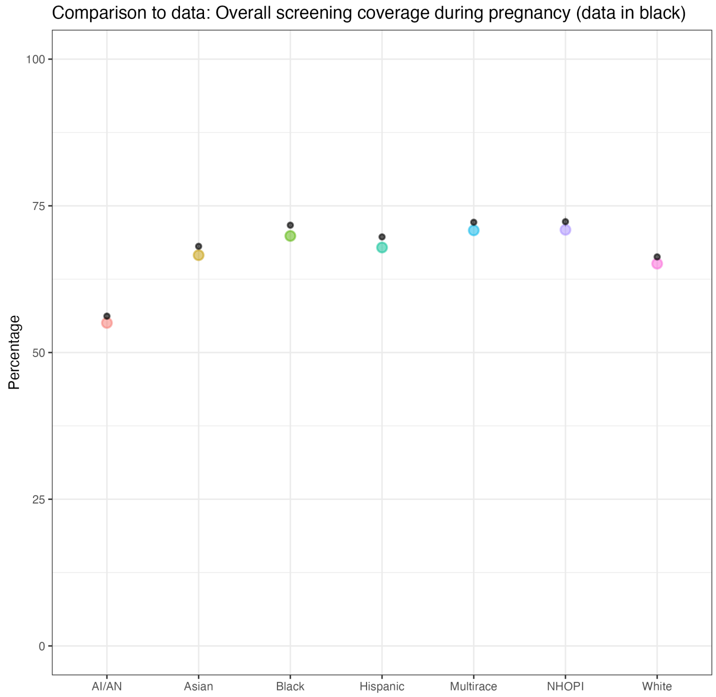

**Figure S5.** Calibration of other congenital syphilis outcomes (excluding stillbirths) against the observed diagnoses in 2019 in the United States among women who are pregnant. [23]

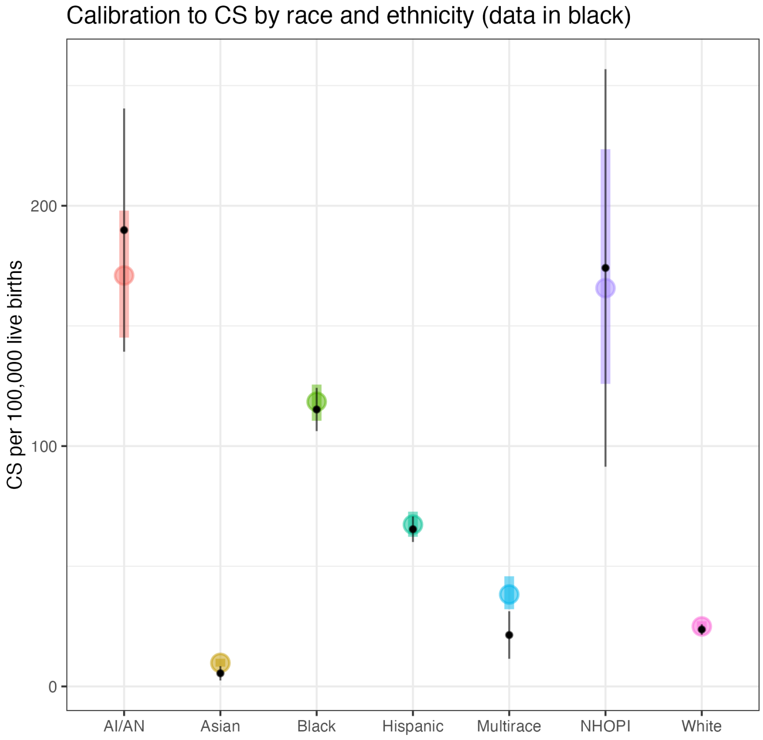

**Figure S6.** Calibration to proportion of other congenital syphilis outcomes (excluding stillbirths) which are diagnosed in women without prenatal care during pregnancy in the United States. Data from 2022. [9]

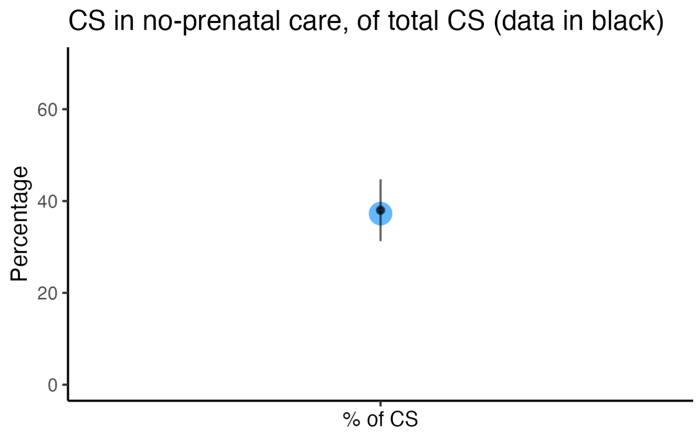

**Figure S7.** Calibration of model to represent data on stillbirth attributable risk per untreated pregnancy excluding the risk of background risk of stillbirth. Data from global meta-analysis. [24]

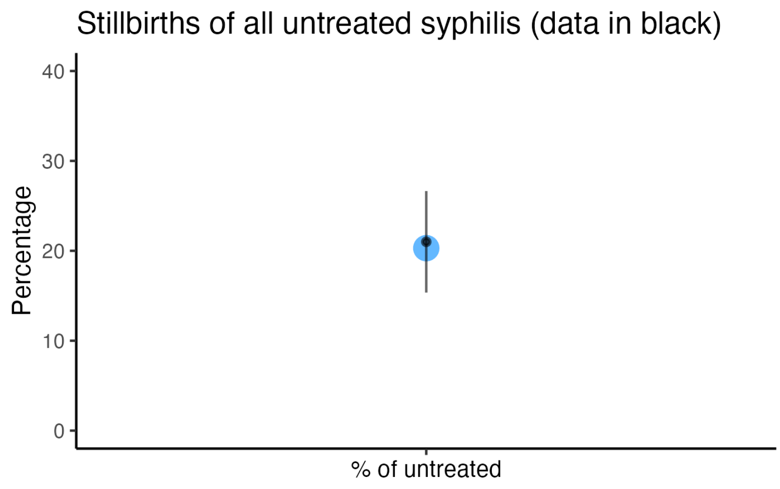

**Figure S8.** Calibration to percentage of pregnant women with syphilis whose infection was acquired while pregnant: Model estimates vs. data from Massachusetts in 2022.*

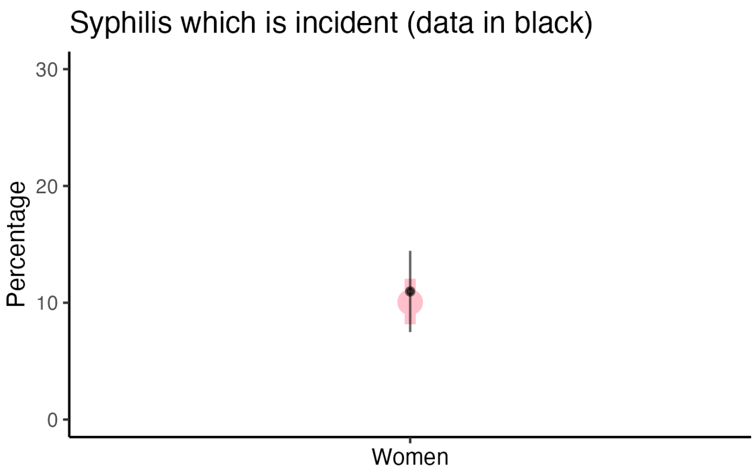

*) Syphilis diagnoses among pregnant women in Massachusetts in 2022 were used to estimate proportion of diagnoses that could have been due to incident infection (based on staging of infection and duration of pregnancy at diagnosis, as well as additional infection available from previous tests when available). Data: Lauren Molotnikov, MDPH.

### Prior and posterior distributions

**Figure S9.** Comparison of model estimated proportion of pregnant women who without prenatal care during pregnancy by race and ethnicity. Compared against prior distribution and the data used to inform the prior (birth certificate data).

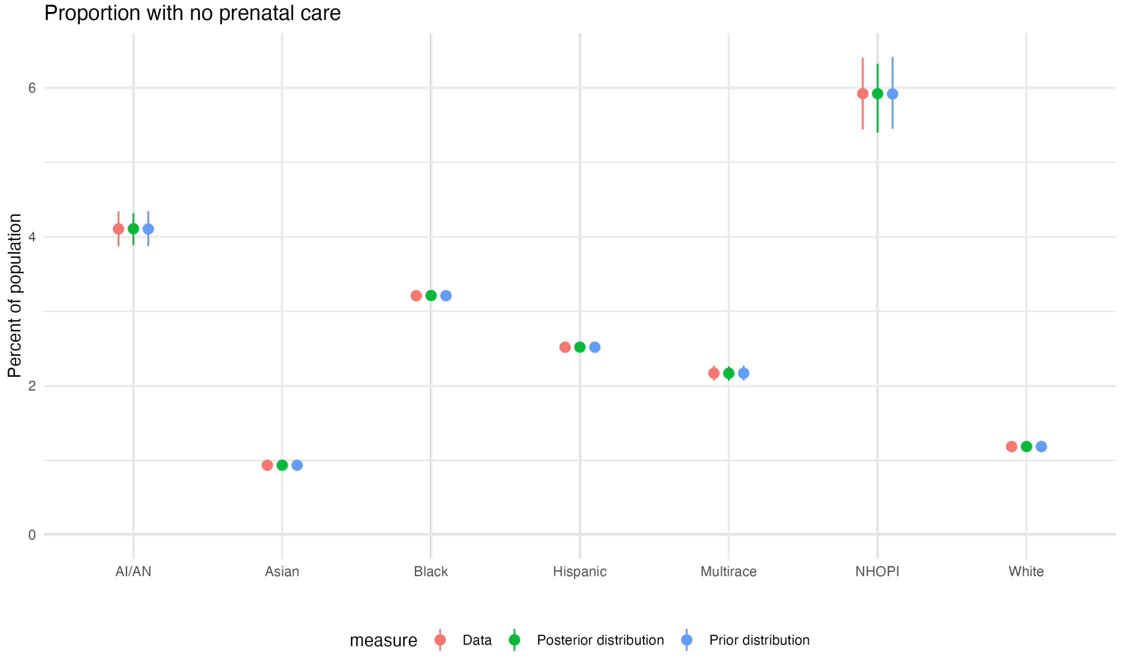

**Figure S10.** Comparison of model estimated prevalence at the beginning of pregnancy. Compared to prior distribution and the positivity in birth certificate data (used to set the upper end of the prior distribution).

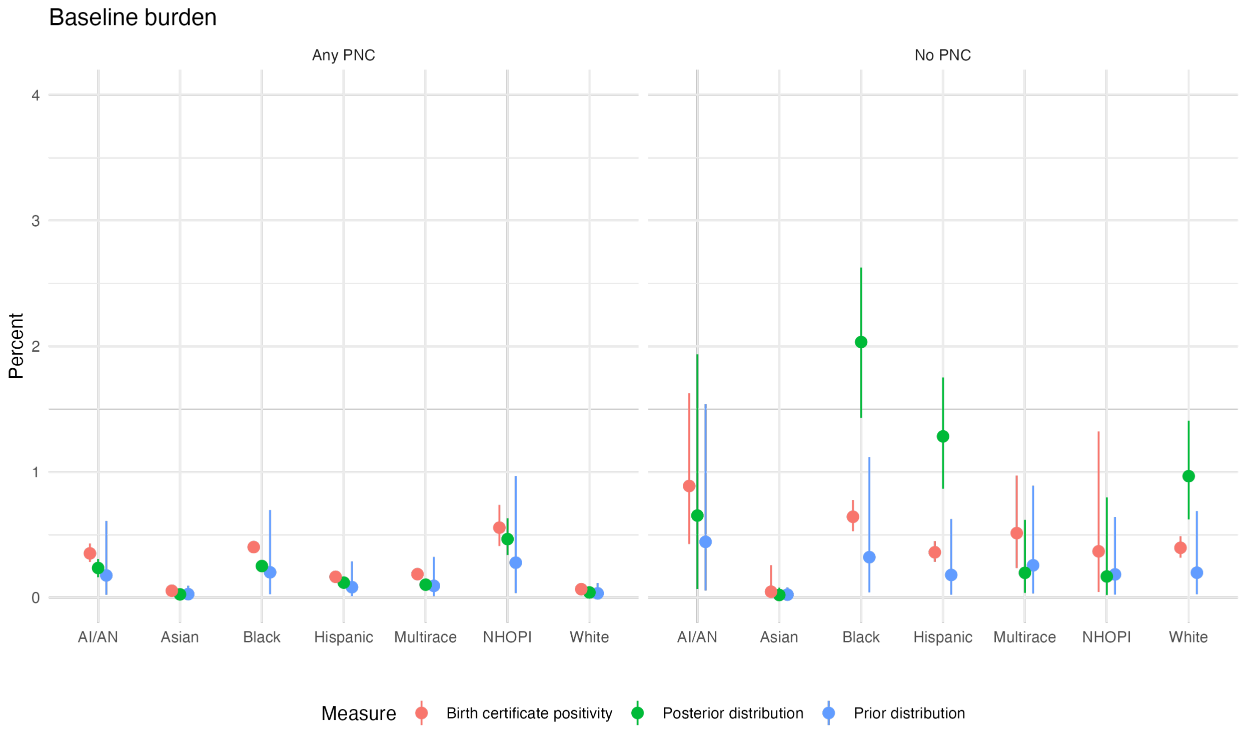
